# Medication and atypical brain maturation in psychosis are associated with long-term cognitive decline and symptom progression

**DOI:** 10.1101/2025.04.02.25325018

**Authors:** Claudio Alemán-Morillo, Natalia García-San-Martín, Richard AI Bethlehem, Lena Dorfschmidt, María Alemany-Navarro, Patricia Segura, Alessia Pasquini, Manuel Muñoz-Caracuel, Manuel Canal Rivero, Jakob Seidlitz, Rosa Ayesa-Arriola, Javier Vázquez-Bourgon, John Suckling, Miguel Ruiz-Veguilla, Benedicto Crespo-Facorro, Rafael Romero-García

## Abstract

**Background:** Clinical progression during psychosis has been closely associated with grey matter abnormalities resulting from atypical brain development. However, the complex interplay between psychopathology and heterogeneous maturational trajectories challenges the identification of neuroanatomical features that anticipate symptomatic decline.

**Aims:** To investigate cortical volume longitudinal deviations in FEP using normative modelling, exploring their relationship with long-term cognitive and symptomatic outcomes, as well as their cytoarchitectural and neurobiological underpinnings.

**Methods:** We collected MRI, cognitive, and symptomatic data from 195 healthy controls and 357 drug-naïve or minimally medicated FEP individuals that were followed up 1,3,5 and 10 years after the first episode (1209 MRI scans and assessments in total). Using normative modelling, we derived subject-specific centile scores for cortical volume to investigate atypical deviations in FEP and their relationship to long-term cognitive and symptomatic deterioration. The resulting centile association maps were further characterized by examining their cytoarchitectural and neurobiological attributes using normative atlases.

**Results:** FEP centiles showed a widespread reduction at treatment initiation, with longitudinal analysis showing an increase during treatment time, indicating convergence toward normal maturation trajectories. Interestingly, this effect was reduced in highly medicated individuals. Additionally, we found that cognitive impairments experienced during early FEP stages worsened under long-term medication. Positive symptomatology was negatively associated with regional centiles, and individuals with higher centiles benefited most from treatment. Cytoarchitectural and neurobiological analyses revealed that regional centiles related to FEP, as well as to symptomatology, were associated with specific molecular features, such as regional serotonin and dopamine receptor densities.

**Conclusions:** Collectively, these findings underscore the potential use of centile-based normative modelling for a better understanding of how atypical cortical development contributes to the long-term clinical progression of neurodevelopmental conditions.

## INTRODUCTION

### Atypical brain structure in FEP

First Episode Psychosis (FEP) is characterised by heterogeneous clinical presentations ^1^, temporal progression ^2^, and responses to medication ^3^. Cross-sectional MRI studies consistently report alterations in frontal and temporal cortices ^4^, with some identifying changes in insular ^5^, parietal ^6^, and occipital regions ^7^. Longitudinal studies have additionally revealed progressive cortical deterioration in FEP, especially in frontal and temporal regions, over 1 ^8^, 2–3 ^9^, 5 ^10^, and 10 years ^11^. Despite these findings, the complex interplay between brain development, sex, and neurodevelopmental pathophysiology complicates the disentanglement of their individual contributions to brain structure and function. Normative modelling addresses this by creating “growth charts” of brain development ^12^, allowing individuals to be ranked and assigned centile scores based on deviations from expected neurotypical trajectories. This approach has been used to identify abnormal structural subtypes in autism ^13^ and deviations in ADHD symptoms ^14^. In psychosis, normative modelling reveals deviations in cortical thickness in temporal areas during high-risk states ^15^ and across the cortex in FEP ^16^, with these deviations diminishing during treatment ^17^.

### Brain structure and clinical outcomes

MRI studies have also extensively explored the clinical presentation of FEP, linking positive symptoms, such as hallucinations and delusions, to thinning in frontal ^18^ and temporal ^19^ cortices. Conversely, negative symptoms, like anhedonia and social withdrawal, are associated with reduced orbitofrontal thickness ^20^. The effects of antipsychotics on brain structure remain debated, with studies reporting both grey matter reductions ^21^ and increased regional thickness ^22^. Cognitive decline in FEP has been observed prior to the onset of psychosis ^23^, while post-onset trajectories vary, with reports of stabilization ^24^, decline ^25^, or improvement ^26^. Normative modelling has shown neurocognitive delays in youth reporting psychotic symptoms ^27^, but the long-term impact of antipsychotics on cognition remains uncertain ^28^.

### Regional vulnerability to atypical development

The spatial patterns of cortical deterioration shared across psychiatric and neurological conditions have been interpreted as a reflection of differential regional vulnerability to pathology ^29^. Under this hypothesis, cortical vulnerability to psychosis-related deterioration may reflect regional differences in cellular composition, neurotransmitter receptors, and metabolism ^29^, with regions of high serotonin and acetylcholine receptor density showing greater atypical deviations ^30^. In this study, we hypothesize that FEP patients exhibit atypical brain development influenced by treatment and medication, which impacts cognitive and symptom progression. We analysed baseline and 10-year follow-up longitudinal data to explore associations between brain deviations, clinical diagnosis, medication, cognition, and symptoms. Using cytoarchitectonic and neurobiological atlases, we additionally characterized psychosis-related brain deviations based on neurotransmitters, cell types, microstructure, and metabolism.

## METODOLOGY

### Subjects

MRI and phenotypic data were collected from healthy controls (n = 195; 120 males; mean age = 29.1 ± 7.63 years) and individuals with first-episode psychosis (FEP; n = 357; 213 males; mean age = 29.8 ± 8.76 years), who were followed longitudinally. Follow-up assessments were conducted at baseline (193 controls, 333 FEP), 1 (62 controls, 96 FEP), 3 (51 controls, 136 FEP), 5 (76 controls, 70 FEP), and 10 (91 controls, 101 FEP) years (See Fig. S1 and S2 for flow diagram for study participants and alluvial diagram of attrition).

Recruitment was conducted through the Program for Attention to the Initial Phases of Psychoses (PAFIP) at Marqués de Valdecilla University Hospital and Valdecilla Health Research Institute (IDIVAL) in Spain. The authors assert that all procedures contributing to this work comply with the ethical standards of the relevant national and institutional committees on human experimentation and with the Helsinki Declaration of 1975, as revised in 2013. All procedures involving human subjects were approved by the CEIC-Cantabria (clinical trial numbers NCT0235832 and NCT02534363) and the Ethic committee of the University of Seville (2024-2534). Following a comprehensive explanation of the study, each participant provided written informed consent prior to enrolment. Participants in the clinical group met DSM-IV criteria for schizophrenia spectrum disorders, including schizophrenia (n=156), schizophreniform disorder (n=99), schizoaffective disorder (n=6), brief psychotic disorder (n=48), and psychosis not otherwise specified (n=24). See a full list of inclusion and exclusion criteria in Supplementary Methods. Detailed demographic and clinical information stratified by sex and diagnosis are provided in Supplemental Material (Figs. S3-S4 and Tables S1-S4).

### Cognitive and symptom assessment

Cognition was assessed for the following domains: (1) verbal memory: Rey Auditory Verbal Learning Test (RAVLT) long-term recall score; (2) visual memory: Rey Complex Figure test (RCFT) long-term recall score; (3) motor dexterity: grooved pegboard (GP), time to complete with dominant hand; (4) executive functions: Trail Making Test part B (TMTb); (5) working memory: WAIS III-Backward Digits (BD) total score; (6) speed of processing: WAIS III-Digit Symbol (DS) standard total score; (7) attention: Continuous Performance Test Degraded-Stimulus (CPT-DS), the total number of correct responses ^31^. For each cognitive test, z-scores were computed by subtracting the mean and dividing by the standard deviation of the control group’s scores. Overall cognitive functioning was calculated as the mean z-score across the seven cognitive tests for each individual. FEP participants were also assessed using the Scale for the Assessment of Negative Symptoms (SANS, 21 items), Scale for the Assessment of Positive Symptoms (SAPS, 30 items), and Brief Psychiatric Rating Scale (BPRS, 24 items).

Principal Component Analysis (PCA) was conducted on cognition and symptom items to compare with average scores used in the calculations. Sensitivity analyses examined (i) the impact of cannabis use and (ii) study dropout on cognitive and symptom measures. Psychological evaluations were performed by experienced psychiatrists using the structured clinical interview for DSM-IV, with diagnoses confirmed at 6 months.

### MRI acquisition and volume extraction

Structural MRI scans were acquired using a 1.5T General Electric SIGMA System (GE, Milwaukee, USA) and a 3T Philips Medical Systems MRI scanner (Achieva, Best, The Netherlands) equipped with an 8-channel head coil. Baseline and follow-up scans for each individual were consistently conducted using the same machine. The imaging protocol included a T1-weighted image that underwent visual inspection for QC. The parameters for 1.5T were: TE=5ms, TR=24ms, NEX=2, rotation angle=45°, FOV=26×19.5cm, slice thickness=1.5mm, voxel size=1.02x1.02x1.5 mm, and a matrix of 256×192, whereas for 3T were TE = 3.7ms, TR=8.2ms, flip angle=8°, acquisition matrix=256×256, voxel size=0.94x0.94x1 mm and 160 contiguous slices. The longitudinal pipeline of FreeSurfer 7.0.0 (recon-all) was used to estimate global cortical volume and regional volume of each region defined in the Desikan–Killiany atlas (34 regions per hemisphere). One regional outlier (>5 SD from the mean) was excluded. Raw regional volumetric data from both scanners were harmonised using ComBatLS preserving age, sex and diagnosis variance ^32^.

### Regional volumetric centile estimation

We leveraged the cross-sectional normative trajectories established by the BrainChart study, which benchmarked cortical volumes from the Desikan-Killiany atlas against data from over 100,000 neurotypical individuals ^12^. Based on these trajectories, we computed out-of-sample centile estimates for our longitudinal sample, using the control cohort to account for scanner-related offsets (https://github.com/brainchart/Lifespan). Following the Bethlehem et al. 2022 pipeline, the volumetric values of left and right homologous regions were averaged to obtain a single measure per region, and centiles were subsequently computed from these averaged values. Age-normed, sex-stratified, and site-corrected deviations were derived and expressed as centile scores. Centile scores represent the relative position of an individual’s grey matter volume with respect to normative lifespan trajectories. Scores below 0.5 indicate volumes lower than the median for age- and sex-matched individuals, whereas scores above 0.5 indicate volumes higher than the median. Residual effects of site or motion (Euler index) on centile scores were assessed using ANOVA.

### Baseline analyses using multiple regression

To evaluate the impact of FEP diagnosis on baseline centiles, a multiple regression was performed for each cortical region using the following model:

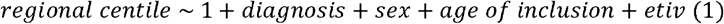

where diagnosis was a binary variable (control vs. FEP), age of inclusion referred to participants’ age at the first scan, and etiv represented cranial volume residuals after age and sex correction (which allowed us to account for anthropometric factors using a single variable). Age of inclusion and etiv were Z-scored prior to regression. All p-values were corrected for multiple comparisons across regions using false discovery rate (FDR).

### Longitudinal analyses using Mixed Modelling

The longitudinal effects of disease and medication on centiles and clinical outcomes were analysed using Mixed Modelling over 1,209 assessments (195 controls, 357 FEP). Antipsychotic medication exposure was assessed at each visit (Fig. S5) and converted into chlorpromazine equivalents (CPZ-equivalents) ^33^.

### Linear Mixed Modelling for predicting regional centiles as a function of disease progression and treatment

Linear Mixed Modelling (LMM) was used to assess the impact of time from the first scan (for controls) and time from treatment initiation (for FEP), collectively referred to as time, in regional centiles. The contribution of age was separated between age of inclusion and time to be able to capture specific interaction effects between treatment time, medication and centiles trajectory in patients:

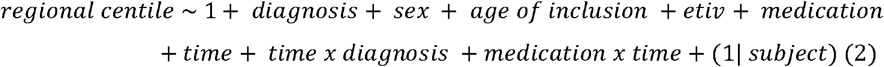

where, in addition to the aforementioned variables, we also considered: medication at the time of MRI acquisition (antipsychotic dose in CPZ-equivalents), time x diagnosis (interaction between time and diagnosis) and medication x time (interaction indicating whether medication alters the effect of treatment time on centiles) and “1|subject” as a random intercept to account for random subject-specific offsets overall. Age of inclusion, etiv, medication and time were Z-scored prior to regression.

Sensitivity analyses repeated these models using raw regional volumes instead of centiles as the dependent variable.

### LMM for predicting cognitive functioning as a function of treatment and regional centiles

After exploring the effects of disease and treatment on centiles, we evaluated how these factors relate to overall cognitive functioning:

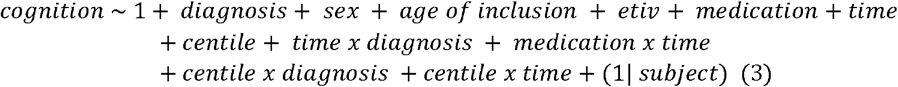

Models were fitted independently for each region, resulting in one parameter estimate per region and covariate. For visualization, a cortical map was generated for each covariate to summarize the regional patterns of association. All p-values were corrected for multiple comparisons across regions using FDR.

### Generalized Mixed Modelling for predicting clinical symptomatology as a function of treatment and regional centiles

Symptomatology (BPRS, SAPS, and SANS scores) was modelled as:

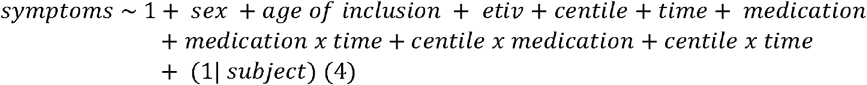

Generalized Mixed Models with a Poisson distribution were used due to the count-like nature of symptom scores and their alignment with exponential decay patterns. Separate analyses were conducted for each region and for each psychometric scale.

Furthermore, for each scale, we conducted a mediation analysis to determine whether the association between medication and symptoms was mediated by the regional centile most strongly associated with symptoms. A bootstrap with 10,000 resamples was used to test for the presence of average direct effect (ADE), average causal mediation effect (ACME) and the total effect (ADE+ACME).

### Cytoarchitectural characterization of centile associations

Cortical centiles associated with phenotypic variables (diagnosis, medication, BPRS, SAPS, SANS) were mapped onto the cortical hierarchies defined by Mesulam atlas (unimodal, paralimbic, idiotypic, heteromodal, https://github.com/ucam-department-of-psychiatry/maps_and_parcs, Table S5, ^34^. ANOVA tests and Tukey post-hoc analyses were used to identify differential distributions of centile associations across Mesulam areas.

### Neurobiological mapping of centile associations

Centile-phenotypic associations were also profiled using 46 molecular and microarchitectural maps derived from MRI/PET studies, expanding on the methodology presented on ^35^ and implemented at https://github.com/netneurolab/neuromaps and https://github.com/RafaelRomeroGarcia/NeurobiologyCentilesPsychosis. These maps included: neurotransmitter receptors (5-HT_1A_, 5-HT_1B_, 5-HT_2A_, 5-HT_4_, 5-HTT, H_3_, D_1_, D_2_, DAT, NET, α_4_β_2_, VAChT, CB_1_, M_1_, MOR, mGluR_5_, NMDA, GABA), cell types (astrocytes, endothelial cells, microglia, oligodendrocytes, oligodendrocytes precursors, excitatory neurons, inhibitory neurons), cortical layers (I-VI), microstructural properties (synapse density, neurotransmitter PC1, gene expression PC1 [which is highly driven by cell types ^36^], cortical thickness, myelin), metabolism (glycolytic index, cerebral blood flow, cerebral blood volume, oxygen metabolism and glucose metabolism), cortical expansion (evolutionary expansion, developmental expansion, allometric scaling from Philadelphia Neurodevelopmental Cohort, allometric scaling from National Institutes of Health). See Table S6 for the full list of neurobiological features included in the analysis.

Principal Component – Canonical Correlation Analysis (PC-CCA; https://github.com/RafaelRomeroGarcia/cca_pls_toolkit) was used to identify combinations of neurobiological features that best explained centile-phenotypic associations ^37^. PC-CCA reduces data dimensionality using PCA to minimize overfitting and then identifies the linear combination (weighted sum of loadings) of the neurobiological maps that best predicts the inter-regional variance of each centiles-phenotypic association map. Statistical significance was evaluated using spatial autocorrelation-preserving permutation tests (spin tests; https://github.com/frantisekvasa/rotate_parcellation) with 10,000 parcellation-specific rotations. To validate the multiple regression patterns identified with PC-CCA, we conducted a sensitivity analysis with Partial Least Squares (PLS). PLS maximizes the covariance between the two variable sets, providing a robust alternative when collinearity might affect the canonical solution ^37^.

## RESULTS

### Impact of diagnosis and treatment on centiles

Using multiple regression on baseline data, FEP patients demonstrated significantly lower total grey matter volume centiles compared to controls (T = -6.20, p <10^-8^), but no reductions in total white matter centiles (T = 1.67, P = 0.095, Table S7; Table S8 for model fit estimates). Consistently, regional analyses showed widespread cortical reductions in centiles associated with diagnosis (**Fig. 1**, Baseline). Covariates such as age of inclusion and etiv correlated positively with centiles, while females exhibited lower centiles (**Fig. 1**, Baseline). Effect sizes and FDR-corrected significance levels are fully provided in the supplementary data. Centiles were not significantly associated with motion artifacts quantified using the Euler index (Spearman’s rho = -0.016, P = 0.587; Fig. S6). As a sensitivity analysis, we recalculated the model including an age of inclusion x diagnosis interaction term, which revealed that the positive effect of age of inclusion on centiles was significantly attenuated in patients (T = – 2.73, P = 0.006).

**Fig. 1.**
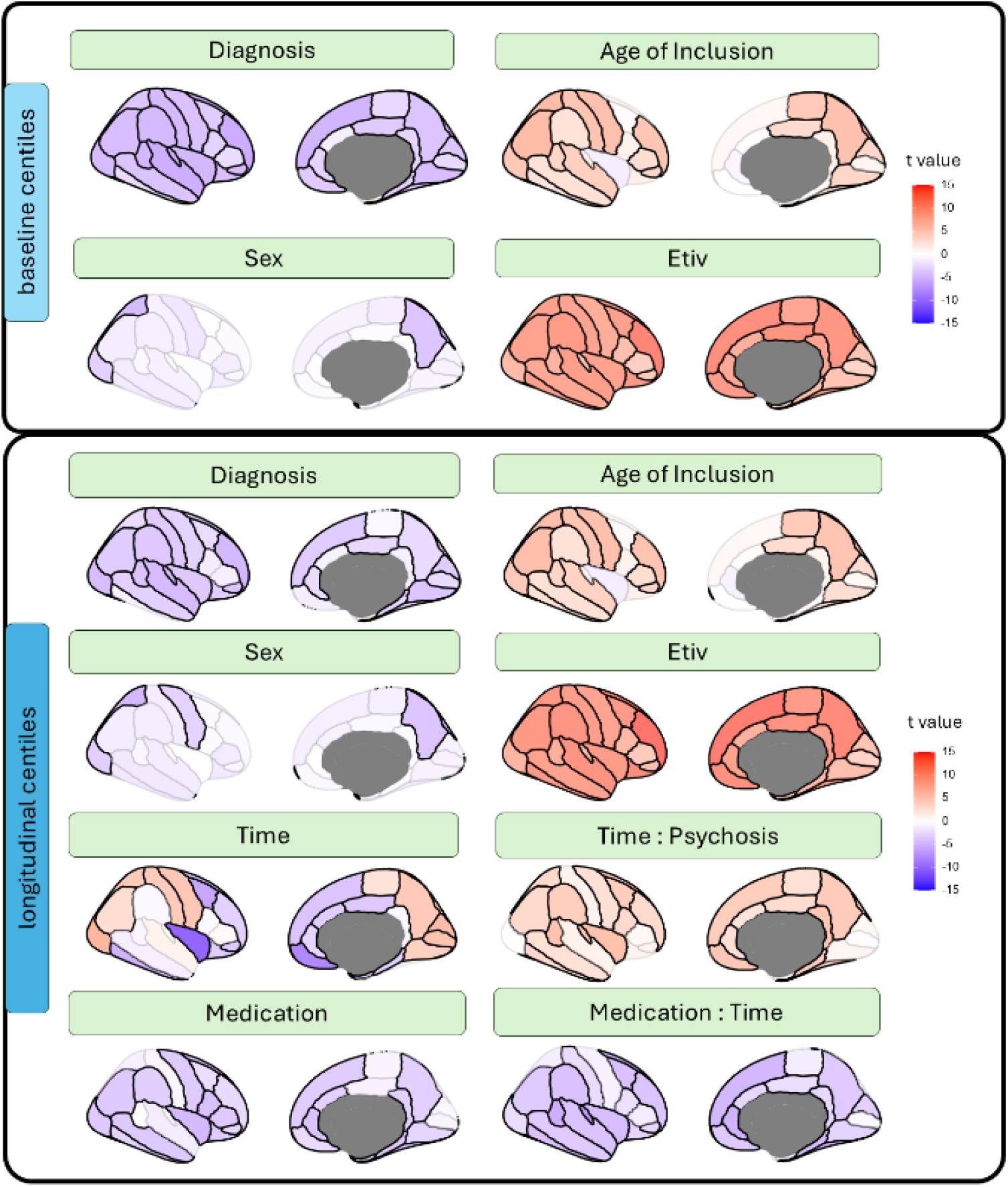
Associations between phenotypic data and covariates with regional centiles. **Top panel**. Contribution of each variable to the multiple regression of baseline data. **Bottom panel** Contribution of each variable to the LMM of longitudinal changes in centiles (up to 10 years follow-up). Scale represents t-values associated with each regression parameter. Non-significant contributions (P_fdr_ ≥0.05) are depicted with desaturated colours.

Longitudinal analyses using LMM over 10 years revealed that longer treatment duration in FEP patients was associated with increased global centile scores, progressively approaching normative values (T = 4.02, p < 10^−4^; Table S9). Regionally, significant diagnosis-centile associations were observed across the whole cortex (**Fig. 1**, longitudinal). The main effect of time showed both positive and negative correlations with centiles; however, the time × FEP interaction revealed a generalized positive association (see Fig. S7A for an illustration of this interaction). Medication (CPZ-equivalent) was negatively associated with centiles, showing also a negative interaction with time. Despite the robust effect of medication, substantial inter-subject variability in centiles persisted between individuals with low versus high medication doses (Fig. S8).

These LMM analyses were repeated using raw volumes instead of centiles. As illustrated in Fig. S9, the principal age- and time-related negative association captured the expected decline in regional volume with aging. Consistent with the centile results, this model also indicated a positive time x FEP interaction, which was attenuated by medication.

### Regional centiles associated with cognitive impairment

Principal component analysis (PCA) of cognitive subscales revealed a first PC explaining 45.3% of the variance, with similar weights across the seven subscales (Table S10), so for simplicity, the mean score was used in subsequent analyses. Cognitive function was negatively associated with FEP diagnosis (**Fig. 2**, diagnosis) but did not correlate with regional centiles (**Fig. 2**, centile - psychosis interaction). Accordingly, the high individual variability limits reliable subgrouping based on centile trajectories (Fig. S10). The observed positive effect of medication, together with the negative medication x time interaction, indicates that the initial therapeutic benefits are attenuated with prolonged treatment (illustrated in Fig. S7B). As a sensitivity analysis, models were repeated by categorizing individuals as low-medicated (CPZ ≤ 300) or highly medicated (CPZ > 300). Interestingly, the global grey matter volume centile showed a weak but significant association with cognitive performance in patients (P=0.04, Table S11). Time had a negative impact on cognition among highly medicated individuals (β_time_-β_time x high-medication_ =0.13 - 0.21 = –0.08; Table S11).

**Fig. 2.**
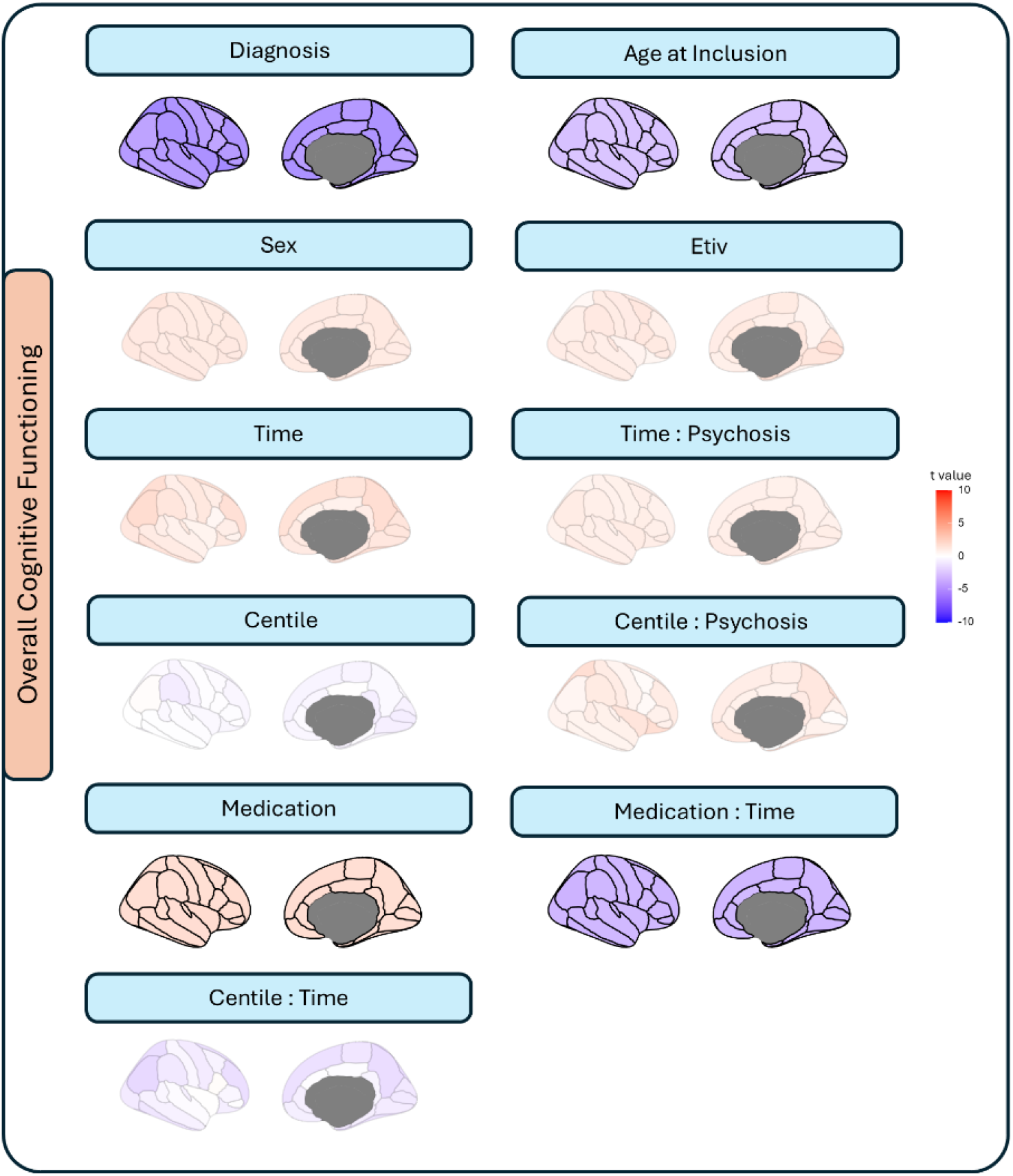
Associations between phenotypic data, covariates and centiles with cognitive performance. Contribution of each variable to the LMM of longitudinal changes in cognition (up to 10 years follow-up). Scale represents t-values associated with each regression parameter. Non-significant contributions (P_fdr_≥0.05) are depicted with desaturated.

### Regional centiles associated with symptomatology

Treatment time correlated negatively with BPRS, SAPS, and SANS scores, indicating symptom reduction over time (**Fig. 3;** see Table S8 for model fit estimates). Medication was associated with reductions in BPRS and SAPS scores but increased SANS scores, likely due to pharmacological specificity for positive symptoms. A positive medication x time interaction indicates that symptom improvement with time was less pronounced in highly medicated patients. Positive symptoms (SAPS) were negatively associated with centiles, particularly overlapping with BPRS, whereas SANS showed a weak correlation with centiles. Younger age at onset was linked to more severe positive symptoms (SAPS). The sensitivity analysis comparing low- versus highly medicated patients revealed that, although attenuated, the effect of time on symptoms remained positive in the highly medicated group (Table S11). No significant differences were observed in cognitive or symptom scores between cannabis users and non-users (Table S12) or between participants who completed the study and those who dropped out (Table S13).

**Fig. 3.**
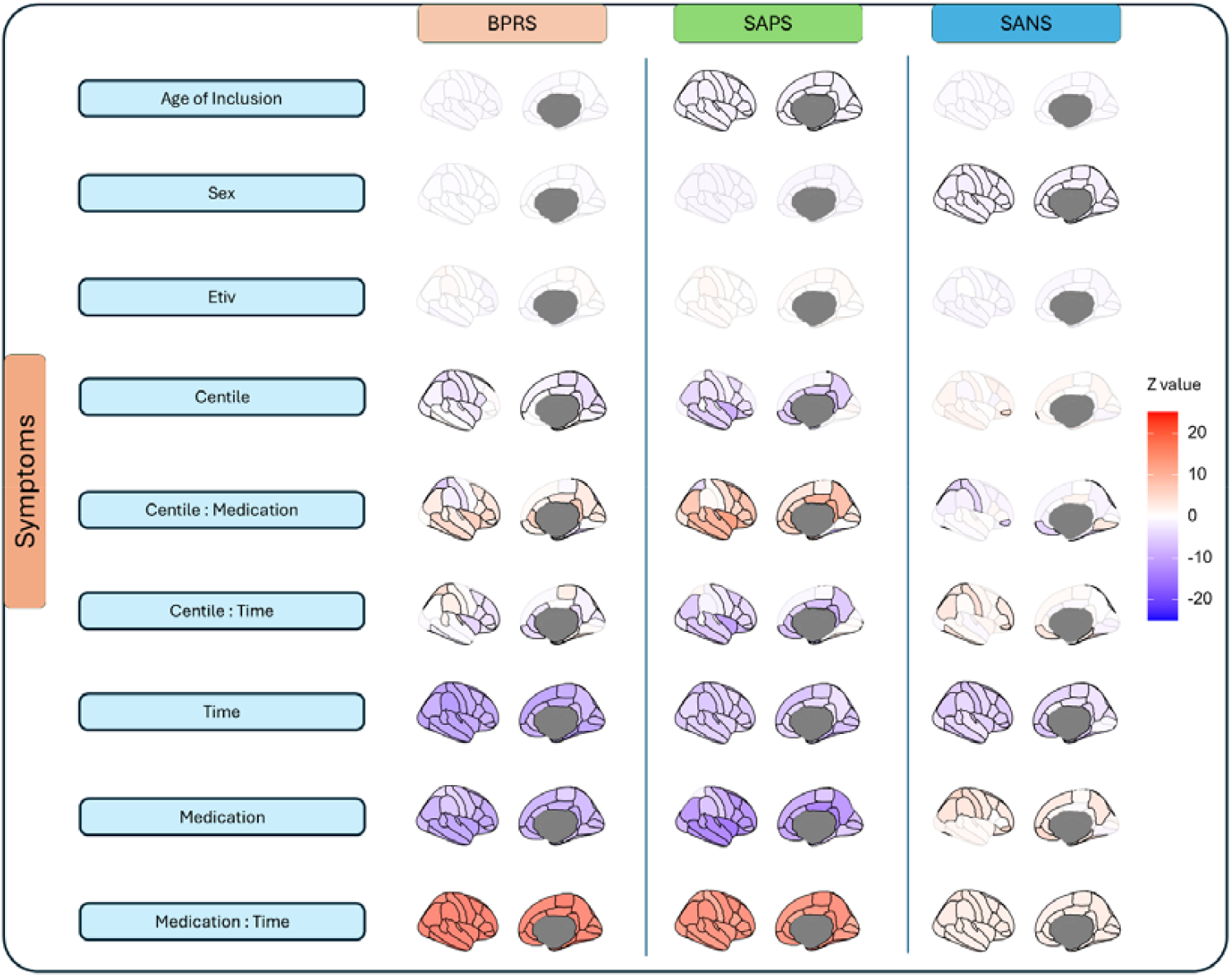
Associations between phenotypic data, covariates and centiles with symptomatology. Contribution of each variable to the GMM of longitudinal changes in BPRS, SAPS and SANS (each column represents different models). Scale represents the Wald z-statistic associated with each regression parameter. Non-significant contributions (P_fdr_≥0.05) are depicted with desaturated colours.

Additionally, mediation analyses were performed to examine the potential mediating effects of centiles and time on the association between medication and symptom severity. No evidence of centile mediation was observed at treatment initiation; however, significant long-term mediation effects were identified. Specifically, the association between medication and symptoms was fully mediated by the insula for SAPS (β=0.15, CI:[0.04,0.27]) and partially mediated by the pars orbitalis cortex for SANS (β=-0.05, CI:[-0.11,-0.01], Fig. S11).

### Cytoarchitectural and neurobiological characterization of FEP-related regional centiles

Using the Mesulam cytoarchitectonic atlas ^34^, we characterized regional associations between centiles, cognition, and symptoms. Centiles linked to BPRS or SANS did not show a preferential topographic Mesulam distribution. However, SAPS displayed significantly stronger negative associations in paralimbic regions compared with idiotypic areas (ANOVA, Post-hoc Tukey, P_fdr_ <0.05, **Fig. 4A**). PCA-CCA was used to capture associations between neurobiological data from normative individuals ^35^ and centiles related to clinical outcomes. Neurobiological mapping revealed that regions associated with FEP diagnosis (Fig. 1) co-located with areas rich in neurotransmitters such as 5-HT_1B_, 5-HT_2A_, 5-HT_6_, as well as neurons and metabolic features (P=0.004, **Fig. 4B**, negative loadings). Conversely, FEP diagnosis-related regions were associated with lower glial cell densities and 5-HT_1A_ neurotransmitter levels (positive loadings). Neurobiological mapping for regions negatively impacted by medication was not significant.

**Fig. 4.**
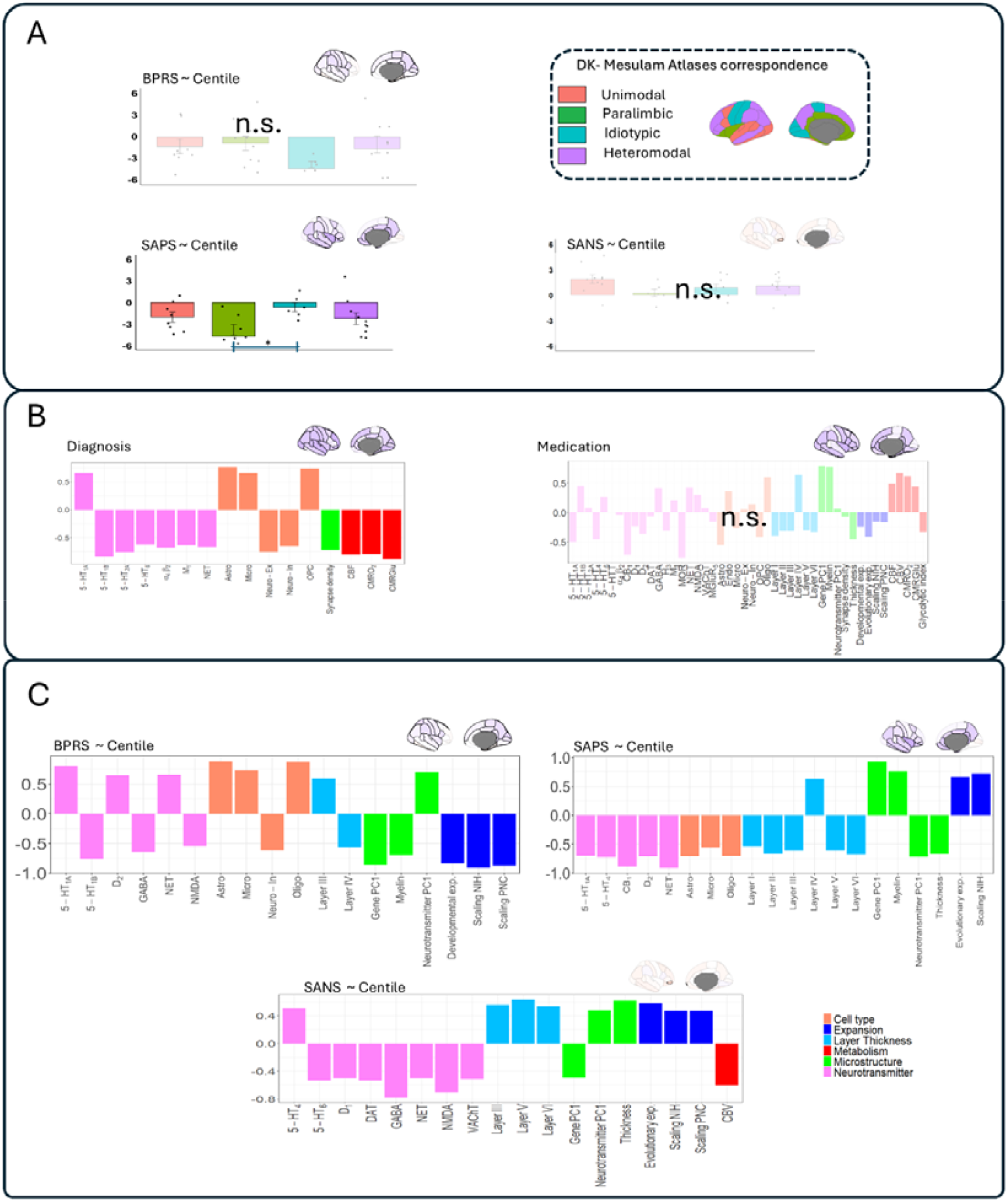
Cytoarchitectural and neurobiological characterisation of FEP-related centiles. **A)** Distribution of t-values shown in Fig. 3 across the four cortices delineated in the atlas of Mesulam. Each dot represents a brain region in the Desikan-Killiany atlas. **B)** Neurobiological maps co-located with the diagnosis-centile and medication-centile associations. Positive loadings indicate neurobiological features that are co-located with regions where centiles were positively associated with the variables of interest (in this case, diagnosis and medication). Conversely, negative loadings indicate co-location with regions negatively associated with the variable of interest. **C)** Neurobiological maps co-located with the association between centiles and clinical outcomes (BPRS SAPS, and SANS). “N.S.” denotes non-significant models (P_fdr_≥0.05). For significant models, only significant (P_fdr_<0.05) loadings are displayed.

Neurobiological features were significantly co-located with the centile-symptoms associations (**Fig. 4C**). Specifically, centiles associated with BPRS co-located with serotonin 5-HT_1A_, D_2_, and norepinephrine transporters, as well as with glial cells (P=0.009). Regions showing negative correlations between centiles and BPRS (i.e. associated with symptom improvement) were co-located with 5-HT_1B_, GABA, NMDA receptors, inhibitory neurons, myelin, developmental expansion and allometric scaling. Centiles associated with SAPS recovery were co-located with 5-HT_1A_, 5-HT_4_, cannabinoid receptors, D_2_, norepinephrine transporters, glia and cortical thickness (P=0.012). Centiles correlated with SANS were co-located with 5-HT_4_, cortical thickness and metabolism (P= 0.01).

Collectively, these results reveal a significant co-location of atypical brain maturation in FEP with specific neurobiological factors, suggesting that they may play an important role in shaping the structural vulnerability to FEP symptomatology. Consistency analyses using PLS instead of PCA-CCA yielded highly similar neurobiological loading estimates (all r-values > 0.78; Fig. S12).

## DISCUSSION

In this study, we examined cortical volume deviations using normative centiles models in both neurotypical and FEP participants scanned over the course of 10 years. Our findings indicated that FEP participants had lower regional centiles than controls in multiple cortical areas, a difference that decreased in intensity over the treatment period. However, this convergence between patients and controls was less pronounced in those who were highly medicated. Psychotic symptomatology was negatively correlated with centiles co-located with regions with high serotonin receptor densities. These results highlight the value of normative approaches for better understanding the long-term relationship between cortical maturation and clinical profiles in psychosis.

Our normative analysis revealed atypical developmental values in FEP patients who exhibited a generalised reduction in regional centile scores ^16^. Previous studies have extensively reported reductions in cortical volume and thickness at different stages of psychosis-related conditions ^5,7,16^. As shown by the ENIGMA consortium, although frontal and temporal cortices are the regions showing the strongest effect sizes when compared to normative controls, schizophrenia is also characterised by a generalised reduction over the whole cortex ^38^. Nevertheless, a major challenge in traditional case-control studies is the lack of personalised metrics capable of detecting individuals with atypical neurodevelopment. Conversely, normative modelling leverages data from thousands of individuals for identifying participants with abnormal trajectories. Using this approach, Worker et al. 2023 ^16^, demonstrated that a large and widespread number of extreme negative deviations in cortical thickness centiles among FEP exist, revealing that this condition is characterised not only by a diminished group-average cortical thickness but also by specific individuals that exhibit severe thickness reductions.

The combination of normative modelling with long-term longitudinal MRI revealed that centiles increased during the treatment period of FEP participants. These findings indicate that grey matter reductions are more pronounced during the early stages of psychosis ^39^, whereas the age-related volume decreases observed in controls are less pronounced in late psychosis ^22^. Berthet et al. (2025) ^17^ also reported this trend toward a reduced difference over time between controls and FEP patients when investigating cortical thickness using normative modelling. Since this study did not include untreated patients, we cannot disentangle the specific effects of treatment from the natural course of the disease and potential late effects of neurodegenerative pathogenesis ^40^. Medication has been previously associated with grey matter loss ^41^, specifically attributed to caspase-3-mediated neurotoxicity ^42^. In this line, we also found that high medication doses were associated with generalized cortical reduction. As suggested by Fusar-Poli et al. 2013 metanalysis ^43^, long-term GM reductions may be the consequence of the cumulative exposure to the antipsychotics, a finding also supported by other studies ^21^.

While describing the anatomical alterations related to the progression and treatment of psychosis is of great interest, understanding how these changes impact patients’ cognitive functionality is of crucial clinical importance. We reported reduced cognitive abilities in drug-naïve and minimally medicated FEP participants immediately after the onset, suggesting that impairment occurs even before the first episode ^23^. Cognitive functionality also improved throughout the treatment, indicating that while medication targets positive symptoms, it may also lead to cognitive improvement, albeit as an indirect effect. ^23^. Previous studies have also demonstrated cognitive improvement in patients after one ^44^ or three ^45^ years of antipsychotic treatment. Nevertheless, our LMM additionally found that medication influenced the association between treatment duration and cognitive function, leading to an attenuation of cognitive recovery over time for patients with higher medication doses. In line with this observation, Kawai et al. 2006 ^46^ demonstrated that dose-reductions lead to improvements in cognitive performance in a limited sample of individuals treated with multiple conventional antipsychotics.

Unsurprisingly, treatment yielded a positive effect on FEP-related symptomatology. As expected, both time and medication primarily had an impact on positive symptoms. Conversely, time had a positive but weaker effect on the reduction of negative symptoms. Despite the fact that medication, especially first-generation antipsychotics, does not alleviate negative symptoms, meta-analyses of placebo-control trials have indeed revealed that most treatments can help reduce negative symptomatology ^47^. Nevertheless, here, we observed opposite trends in the model slopes of time (negative) and medication (positive) with SANS, indicating that heavily medicated individuals do not experience negative symptoms improvements over time. This may reflect the diverse and distinct recovery trajectories of illness, with a significant proportion of patients experiencing relapse and showing no improvement in negative symptoms ^48^. FEP symptomatology was also associated with regional volumes, with higher centiles values being associated with lower positive symptoms. Similarly, previous studies have associated positive symptoms with cortical alterations in prefrontal, temporal and parietal areas ^19^. Conversely, negative symptoms exhibited a weak correlation with centiles, suggesting that while treatment and medication may provide moderate improvement in this domain, these symptoms remain relatively stable and do not significantly influence cortical plasticity. However, further placebo-controlled trials will be necessary to elucidate this ^48^.

Neurobiologically, serotonin neurotransmitters were predominantly present in regions associated with psychosis diagnosis. This may be related with the role of this neurotransmitter mediating the effect of antipsychotic medication which affects mood and cognitive function ^49^. We also reported differences in centile associations across receptor subtypes such as 5HT_1A_ and 5HT_2A_ which could be a reflection of its complex role in psychosis ^50^. Additionally, the co-location of dopamine receptors in regions associated with symptoms suggests a potential pathway whereby elevated levels of antipsychotic medications disproportionately affect dopaminergic regions. Moreover, diagnosis and symptom-related centiles were not only associated with neuronal density maps but also with glial cells, indicating a potential involvement of immunological and inflammatory risk factors ^51^.

### Limitations

Several limitations must be acknowledged. Firstly, the primary inclusion criterion was the diagnosis of FEP at baseline, regardless of subsequent follow-up diagnoses, which predominantly included schizophrenia but also other psychotic disorders. Secondly, the use of CPZ-equivalents provides a simplified representation of the heterogeneous antipsychotic treatments across the sample, allowing us to capture the overall medication effect with a single parameter. However, different classes of antipsychotics may have differential effects on centiles and clinical outcomes. Including all drug-specific doses was not feasible, as this would substantially increase the number of variables and the risk of overfitting. Finally, higher medication doses were generally prescribed to individuals with more resistant positive symptoms. However, these prescriptions were influenced by factors such as age of onset and treatment duration, complicating the disentanglement of individual contributions in the models.

This brain-wide association study (BWAS) demonstrates that centiles are associated with clinical outcomes in FEP. Nevertheless, growing evidence highlights important concerns: BWAS are susceptible to inflated effect sizes and low replication rates unless samples include thousands of individuals ^52^. Recent findings by Kang et al. 2024, have shown, however, that longitudinal designs can substantially improve replicability ^53^. In this study, participants were comprehensively assessed over a 10-year period. While replication gains plateau after the first follow-up, particularly for structural markers, which exhibit limited within-subject variability, longitudinal assessments remain valuable for capturing the pronounced phenotypic decline in cognition and symptoms over time in FEP. Importantly, this high within-subject phenotypic variability is critical for enhancing replicability, especially when modelled separately from between-subject variability ^53^, as implemented here.

We aimed to maintain the models as parsimonious as possible. With 552 participants, up to 11 fixed variables and at least 449 degrees of freedom, the resulting balance was appropriate for robust and replicable estimation of model parameters ^54^. Nevertheless, aspects such as the inclusion of moderate covariates, potential underlying collinearity, or missing data further contribute to the complexity of assessing the actual statistical power and the potential risk of overfitting of the models.

Centiles derived from cross-sectional brain charts have demonstrated to underestimate longitudinal brain changes, resulting in inaccurate predictors of longitudinal individual changes ^55^. In this study, which includes extensive follow-up data, our objective was not to perform individualised age-related predictions, but to identify volumetric deviations associated with long-term cognitive and symptomatic outcomes in FEP. To account for residual age effects retained by the centiles, both age of inclusion and time were included as main effects in the models. However, longitudinal normative modelling was not performed due to the limited sample size. Future work will benefit from calibrating brain reference charts with longitudinal datasets ^55^. This will be crucial for characterizing individual-level trajectories and for identifying subgroups that may account for clinical heterogeneity and differences in treatment response.

## Conclusions

This study applied a normative modelling approach to demonstrate that FEP is characterised by regional decreases in cortical centiles. Centiles recovered to normative values during treatment but they were negatively affected by high doses of long-term medication. Similarly, improvements in cognitive abilities and symptomatology were positively associated with time, although were negatively influenced by prolonged medication use. Regions sensitive to symptomatology co-located with higher serotonin receptor densities and other neurobiological features. Collectively, these findings provide critical insights into the long-term interactions between cortical structure, treatment, medication, cognitive function, and symptom profiles.

## Data and Code availability

All code and non-clinical data used to perform the analyses can be found at https://github.com/RafaelRomeroGarcia/LongitudinalCentilesFEP/. Data from patients is available upon request in accordance with ethical regulations. All statistical results are provided in a supplementary Excel file.

## Acknowledgements

We thank Agoston Mihalik, Golia Shafiei, Bratislav Misic, Victor Ortiz and Lifespan Brain Chart Consortium for their contribution.

## Author Contributions

C.A-M performed data curation, methodological design, data analysis, and drafted the manuscript; N.G-S-M, R.A.I.B, L.D, M.A-N, P.S, A.P, M.M-C, M.C R, J.S, R.A-A, J.V-B, J.S, M.R-V, B.C-F and R.R-G contributed to data acquisition, provided advice on data analysis, and participated in writing and editing the manuscript. R.R.G. also contributed to conceptualization, first manuscript draft, and supervision of the work. All authors approved the submitted version of the manuscript.

