## Supplemental Material for "Medication and atypical brain maturation in psychosis are associated with long-term cognitive decline and symptom progression"

*Supplementary Material*

### Supplementary Methods

#### Subjects

PAFIP (Programa de Atención a las Fases Iniciales de Psicosis) participants underwent screening based on the following criteria[[1]](https://www.zotero.org/google-docs/?yxgLDN): (1) age range of 15 to 60 years; (2) meeting DSM-IV criteria for a primary diagnosis of schizophreniform disorder, schizophrenia, schizoaffective disorder, brief reactive psychosis, schizotypal personality disorder, or psychosis not otherwise specified; (3) permanent residency within the designated catchment area; (4) absence of prior exposure to antipsychotic medication or, if previously treated, a cumulative lifetime history of sufficient antipsychotic symptoms of at least moderate severity, as evaluated by one of the five items of the Scale for the Assessment of Positive Symptoms. (SAPS).

Patients were ineligible for inclusion in the study if they met any of the following criteria: 1) met DSM-IV criteria for drug dependence (excluding nicotine dependence), 2) met DSM-IV criteria for mental retardation, or 3) had a history of neurological disease or head injury. There were no additional exclusion criteria for MRI scans except those related to logistical constraints (e.g., claustrophobia, braces). The control group comprising healthy volunteers was recruited from the community via advertisements. They were required to have no current or previous psychiatric disorders, intellectual disability , neurological conditions, or significant medical illnesses, including substance dependence and prolonged loss of consciousness, as assessed using a condensed version of the Comprehensive Assessment of Symptoms and History [[2]](https://www.zotero.org/google-docs/?Pu8DnI). Selection criteria aimed to achieve similar distributions in age, gender, and drug use history as the patient group. Furthermore, clinical records and family interviews were utilized to verify the absence of psychosis in first-degree relatives. Following a comprehensive explanation of the study, each participant provided written informed consent prior to enrolment.

### Supplementary Figures

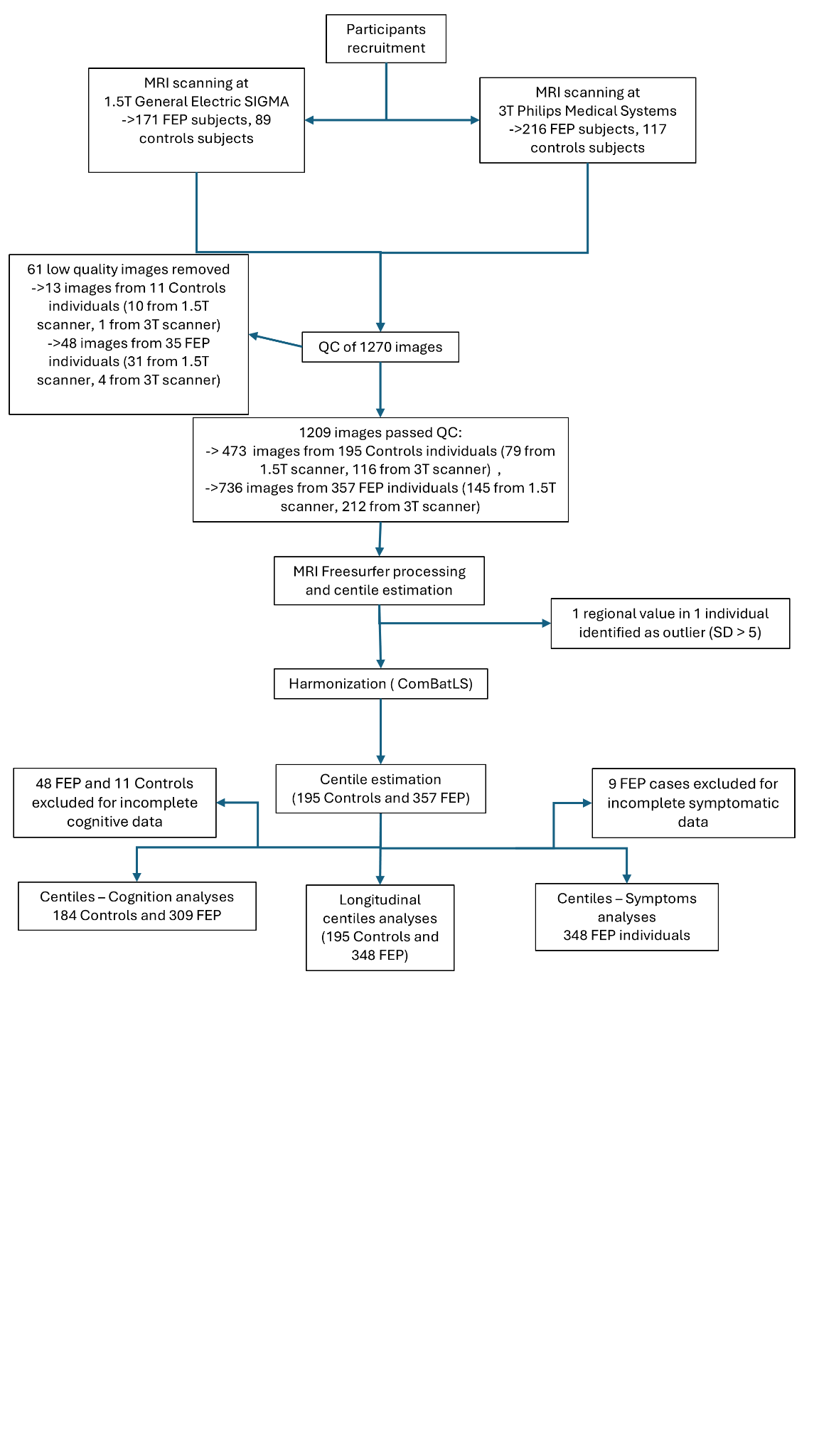

**Fig. S1.** Flow diagram for study participants.

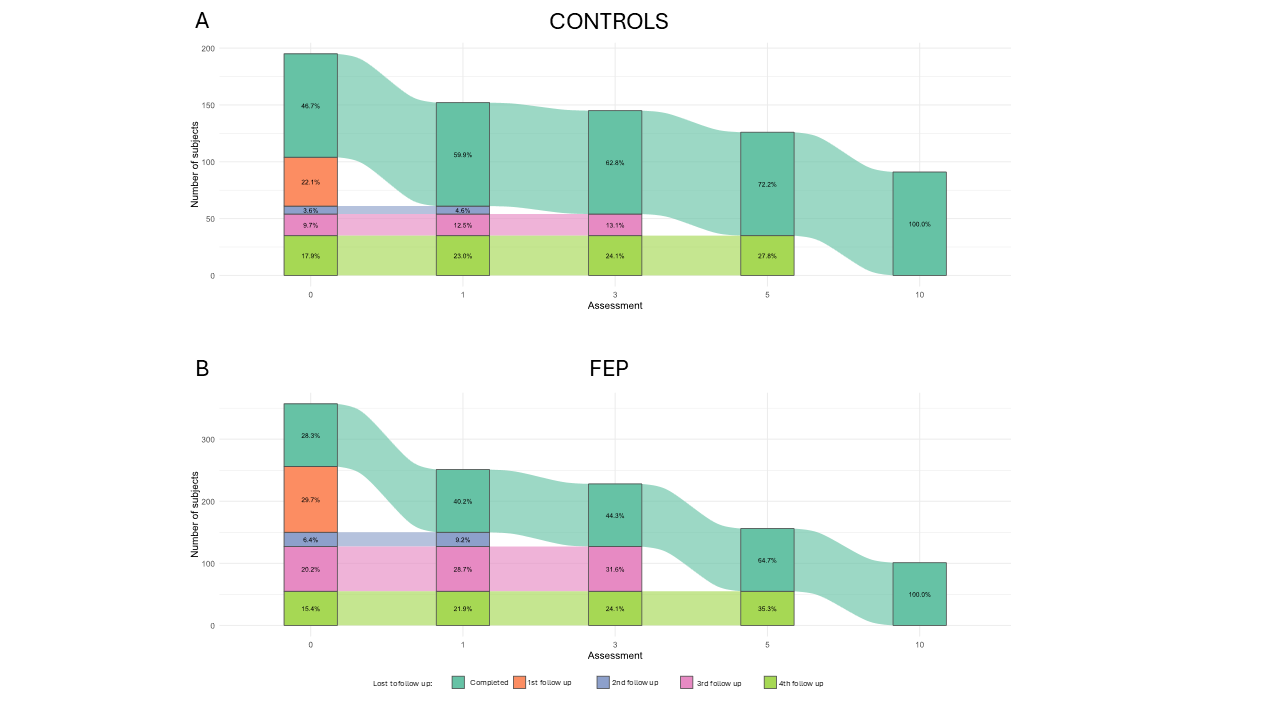

**Fig. S2.** Alluvial diagram illustrating participant attrition.

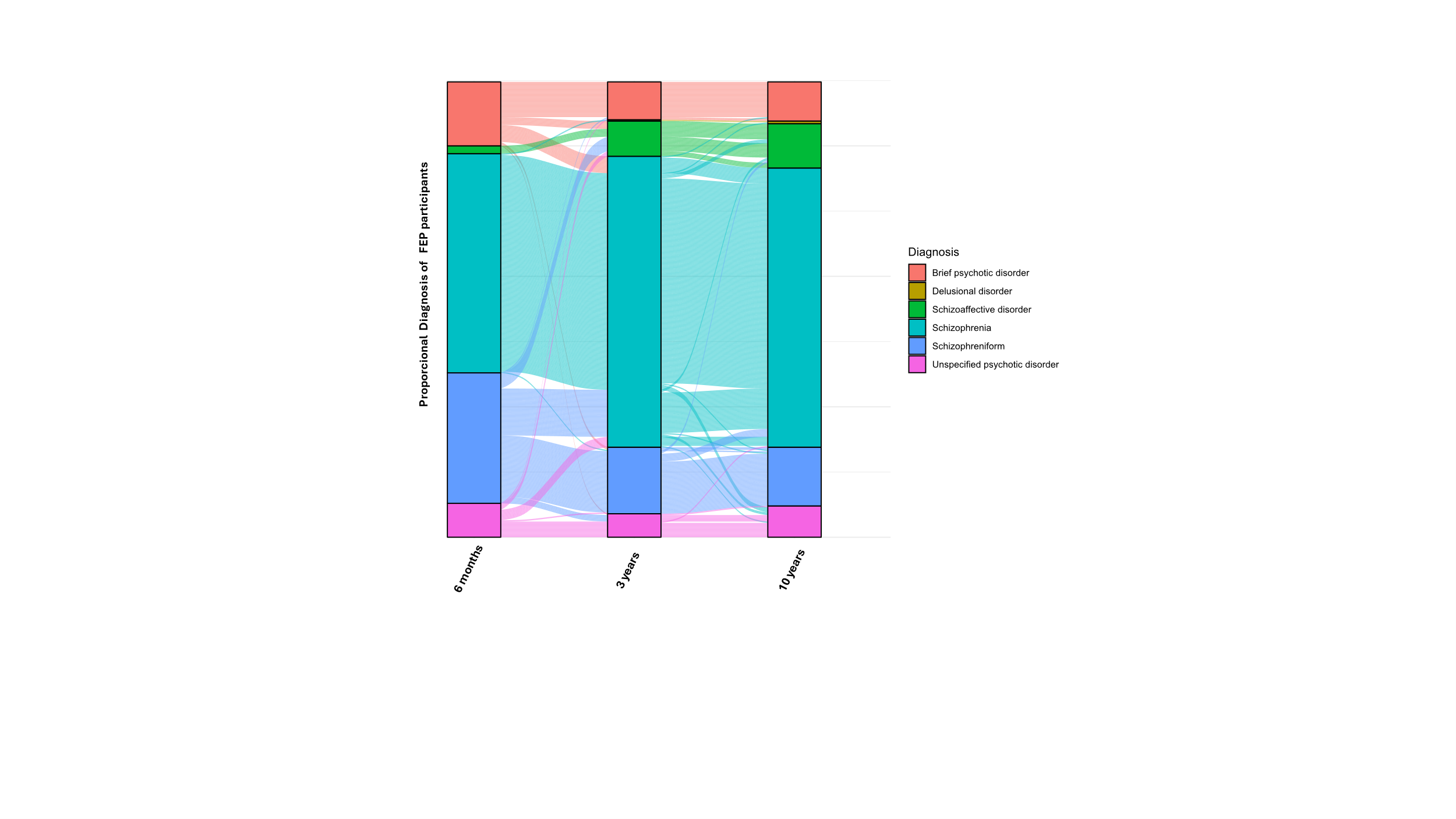

**Fig. S3**. Alluvial plot depicting participant transitions across diagnostic categories over three assessment timepoints. The plot shows stacked bars (strata) representing the distribution of diagnoses at each timepoint and connecting flows (alluvia) indicating the proportion of participants transitioning between diagnostic categories. The height of each stratum and the width of each flow are proportional to the ratio of FEP participants with each diagnosis.

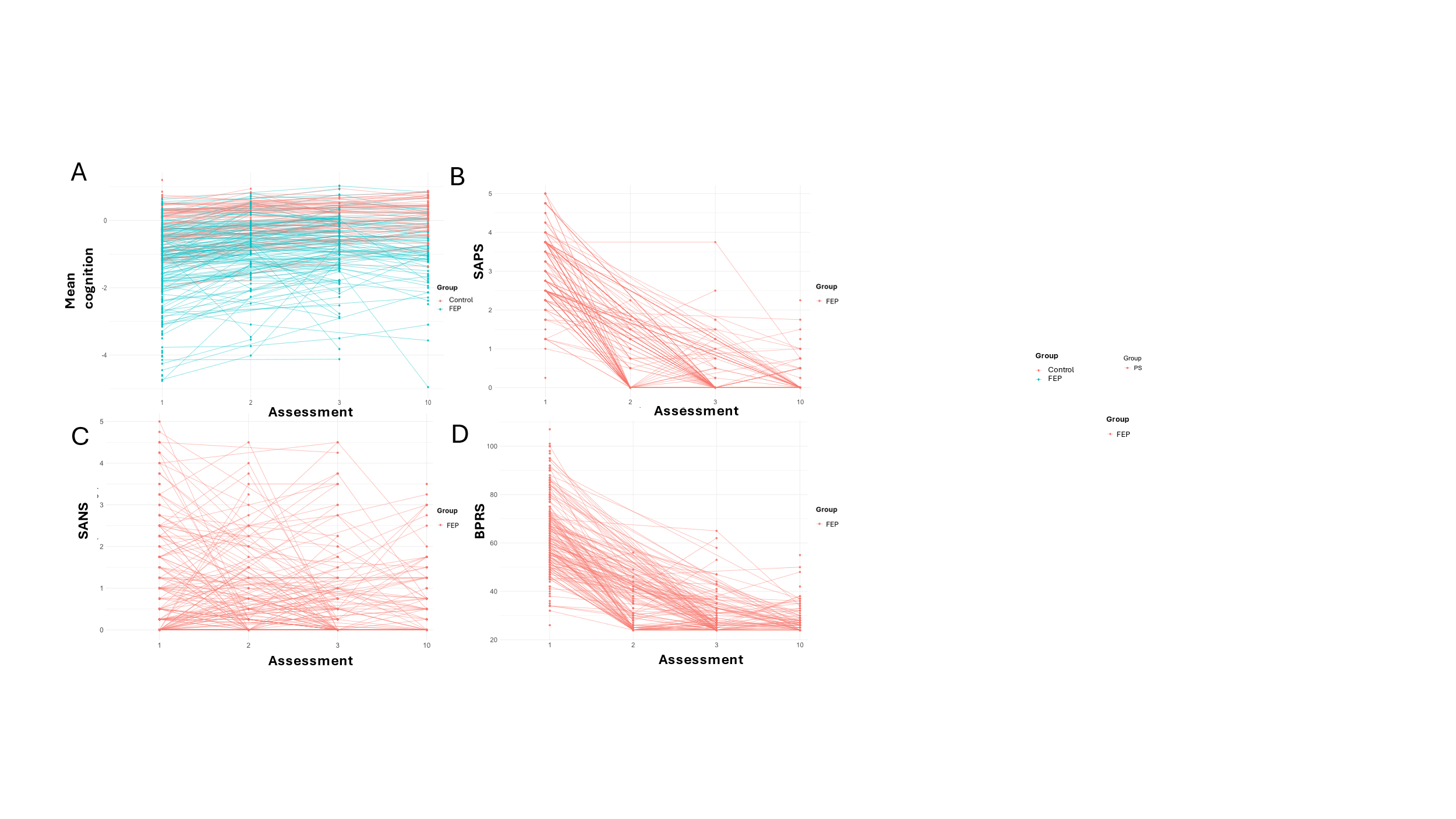

**Fig. S4**. Longitudinal progression of cognition and symptoms in the cohort.

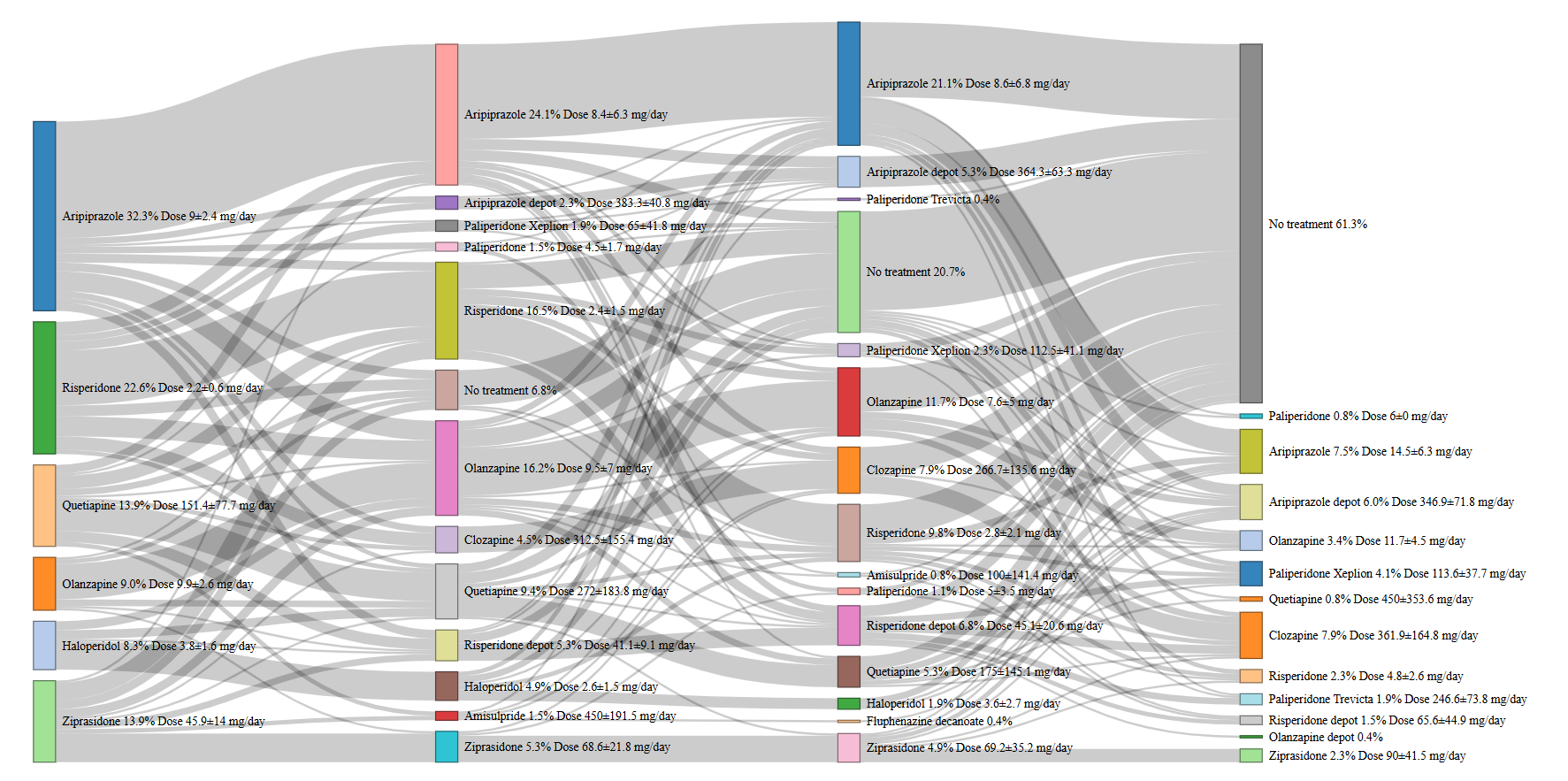

**Fig. S5.** Alluvial diagram illustrating changes in medication over the course of treatment.

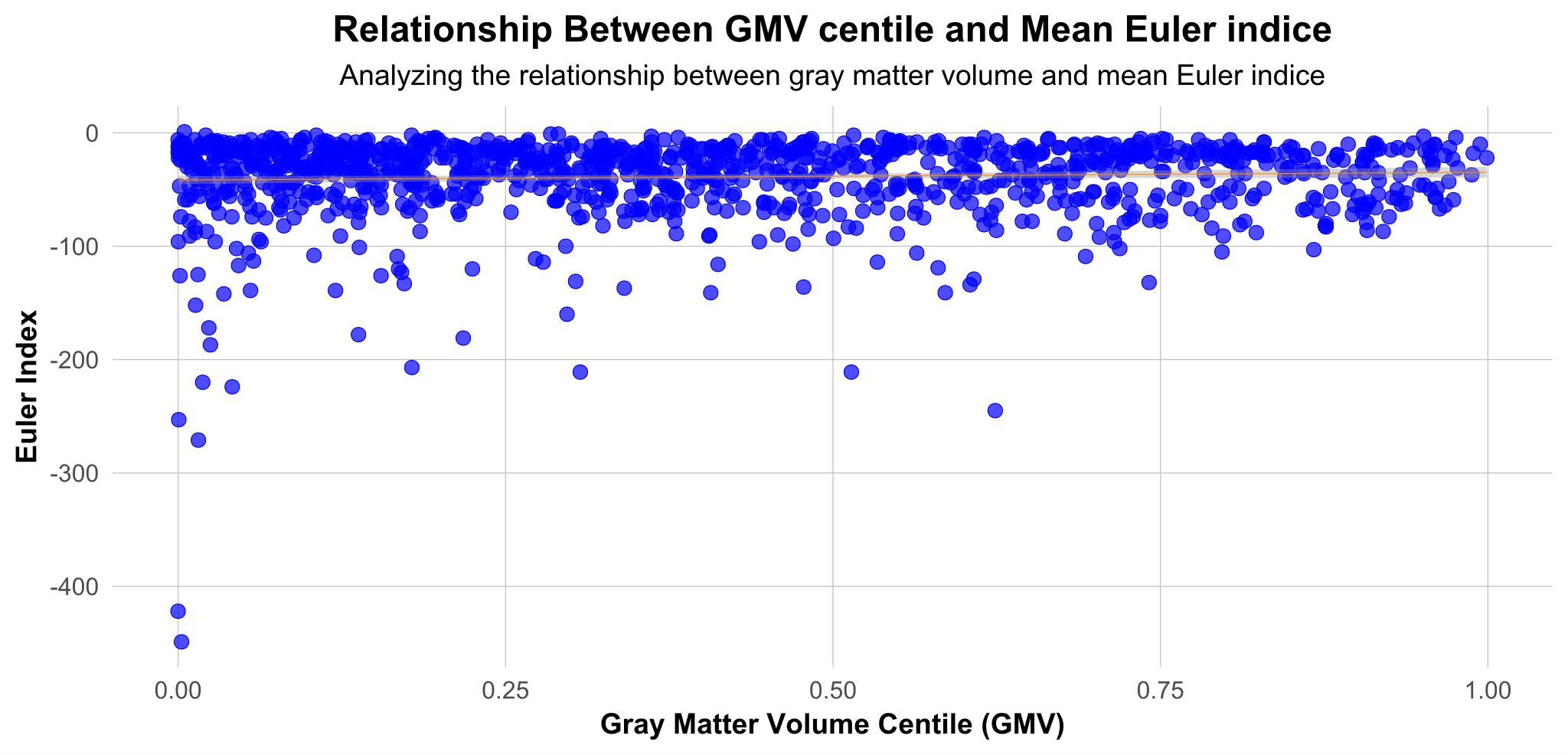

**Fig. S6.** Relationship between the Euler index of each image and total gray matter centile volume.

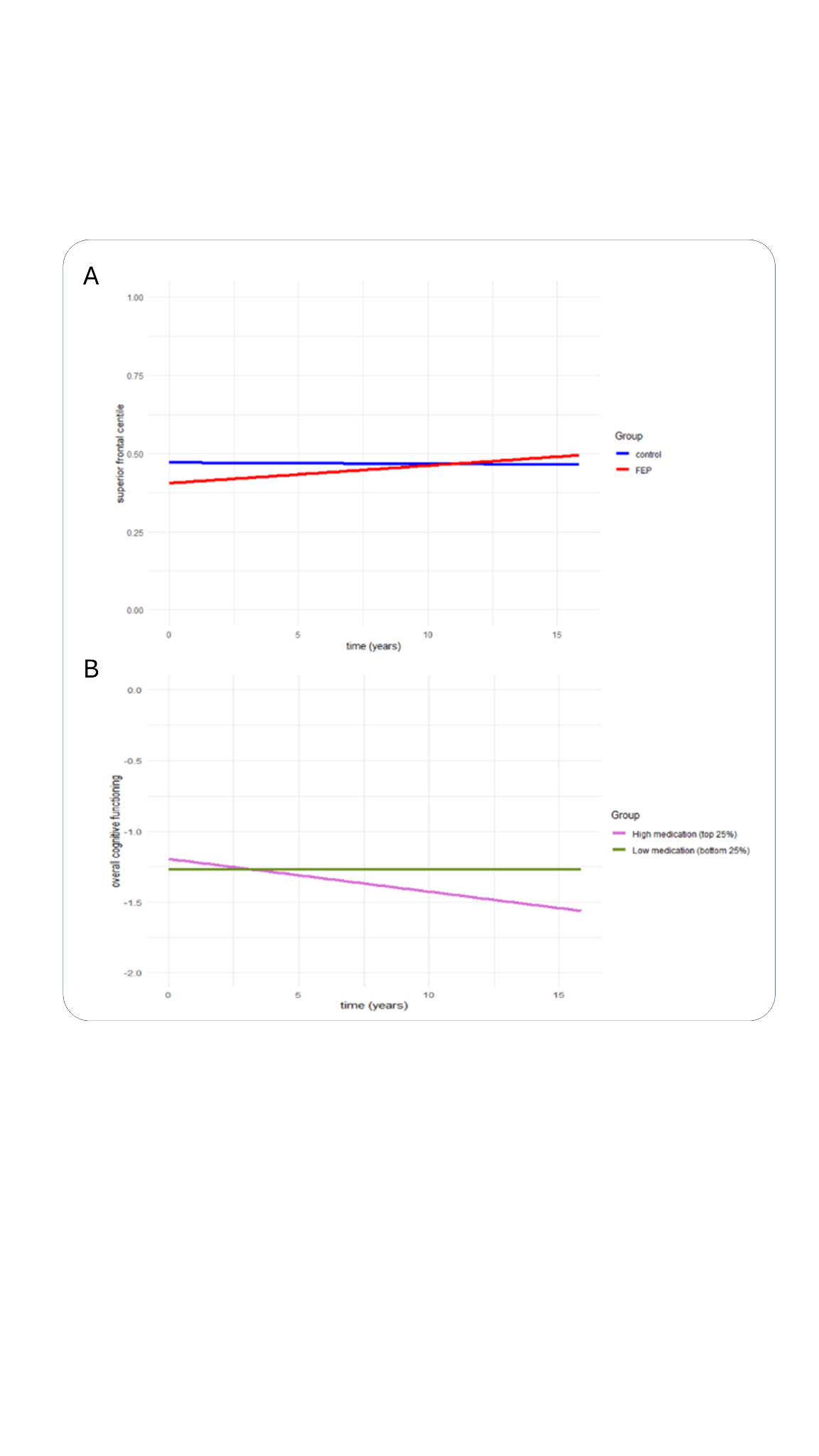

**Fig. S7. Illustrative representation of interaction effects between variables in a representative centile.** (A) Relationship between superior frontal centile and treatment duration, stratified by diagnosis. (B) Relationship between overall cognitive functioning and treatment duration, stratified by medication.

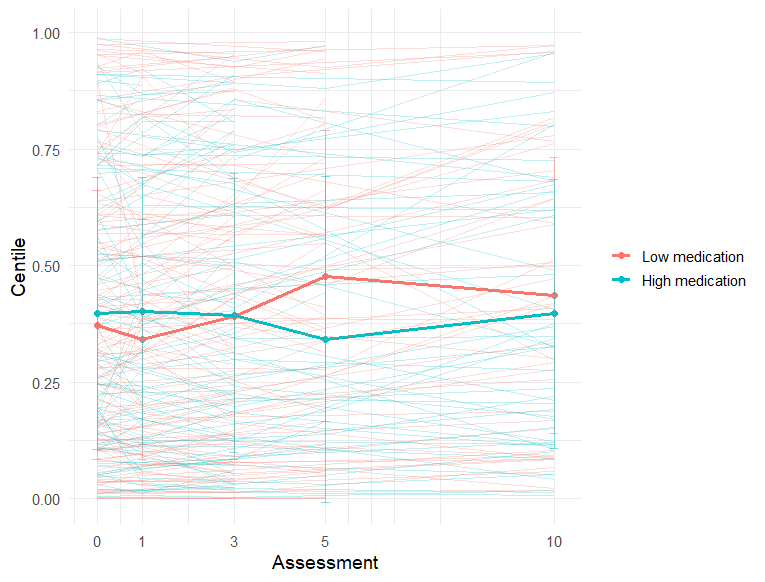

**Fig. S8.** Global centile across assessments color-coded by patients’ medication status. Patients were classified as having high or low medication depending on whether their CPZ-equivalent dosage exceeded 300 at any point during treatment.

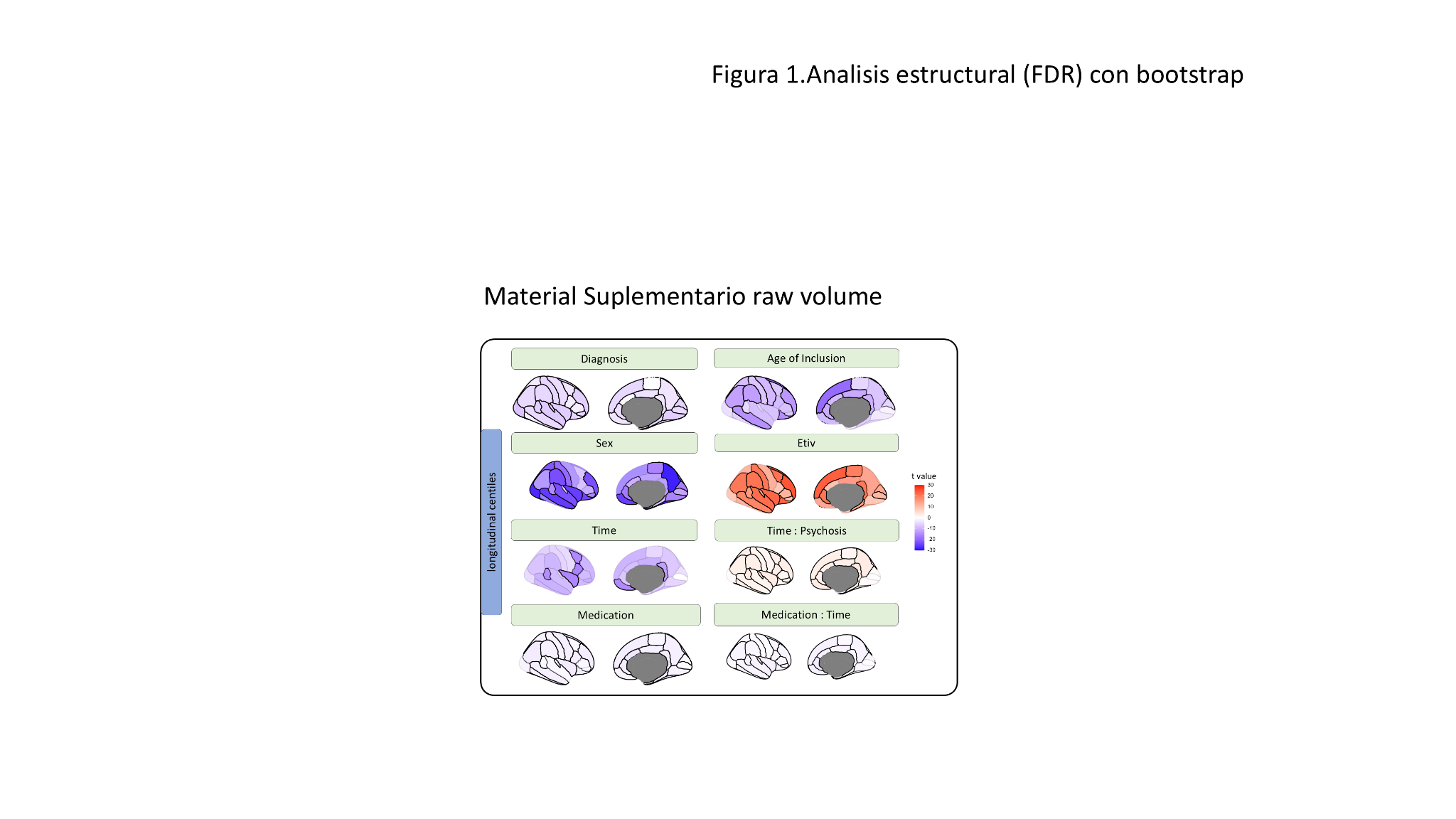

**Fig. S9**. Associations between clinical data and covariates with raw volumes.

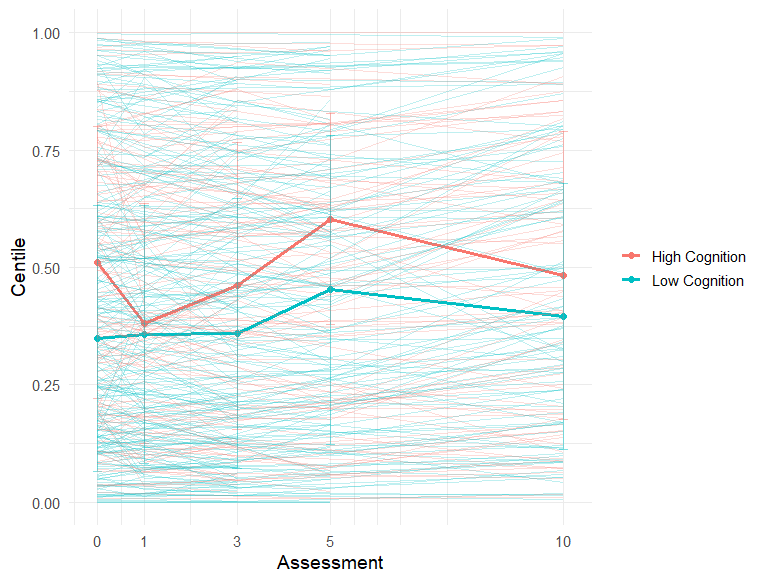

**Fig. S10.** Global centile across assessments color-coded by patients’ final cognitive status, categorized as high or low depending on whether they scored above or below the overall final cognition mean of all study participants.

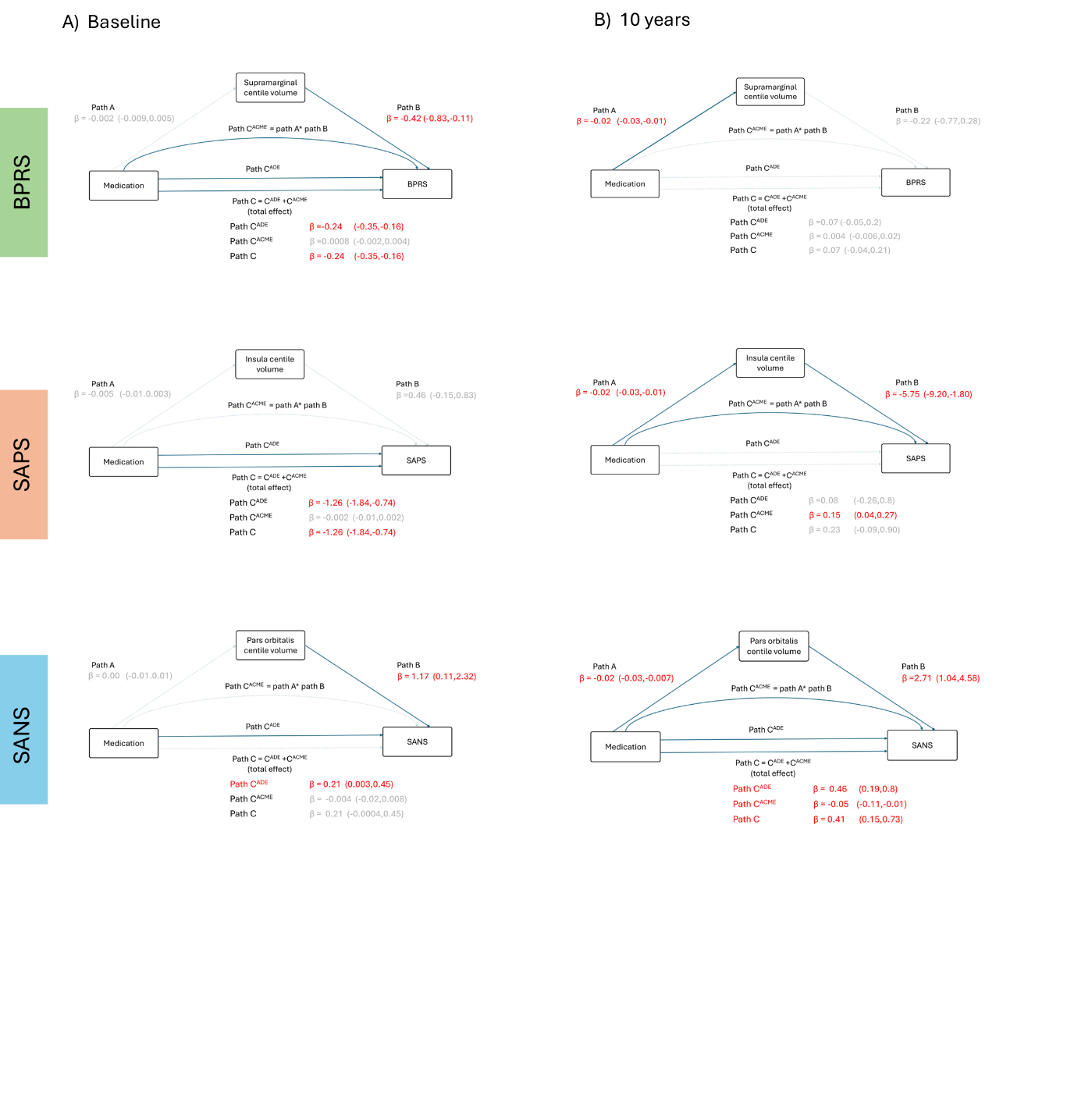

**Fig. S11.** Mediation analyses of centiles and treatment time on the association between medication and symptomatology. (**A**) Mediation effect at treatment initiation (t=0). (**B**) Mediation effect at 10 years follow-up. Dashed lines show no significant paths. The overall mediation effect of path C is the sum of the average direct effect (path C^ADE^) and the average causal mediation effect (path C^ACME^).

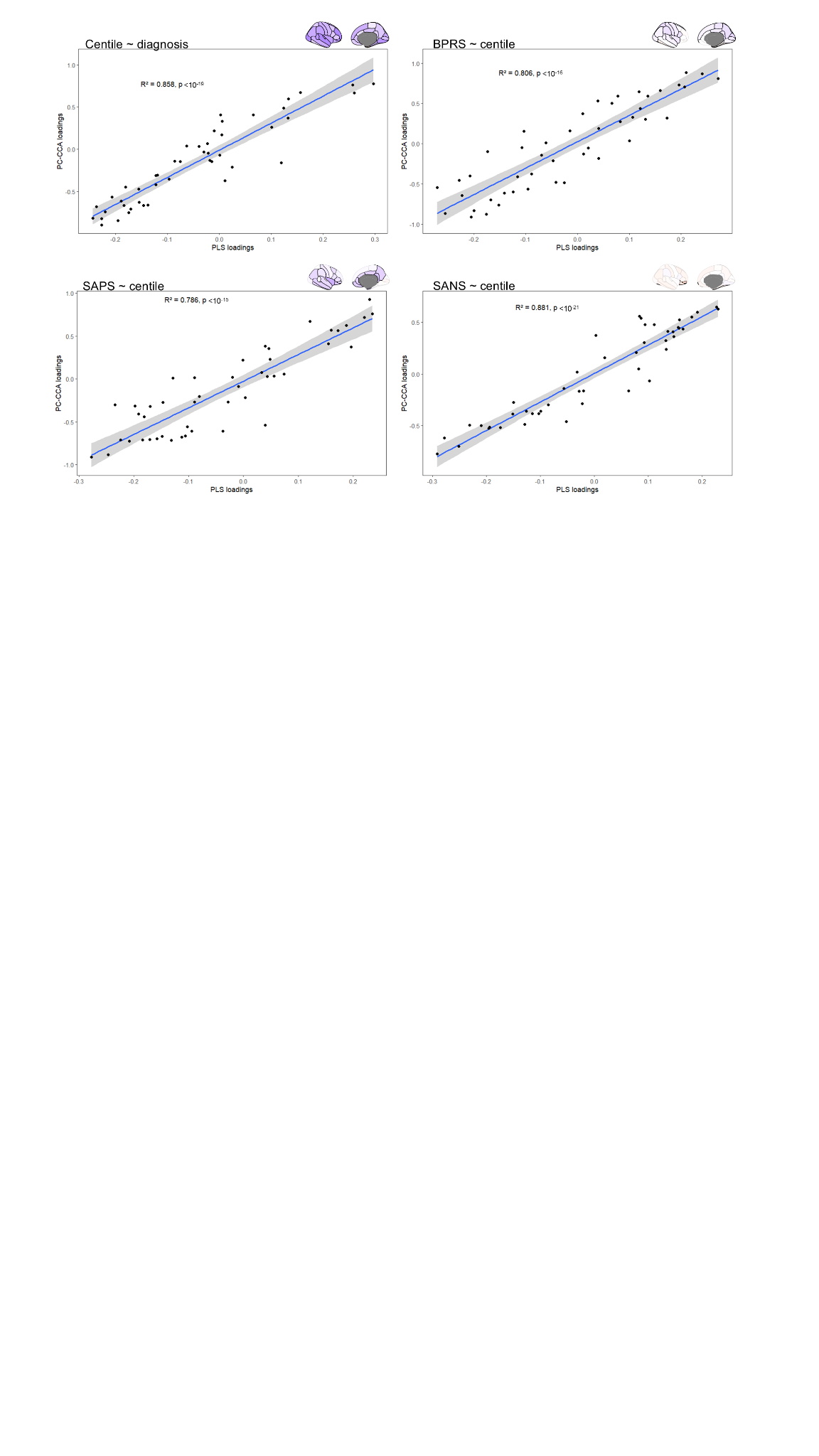

**Fig. S12**. **Correlation between PCA-CCA and PLS loadings.** Scatterplots showing the similarity of PCA-CCA and PLS loadings derived from mapping neurobiological features into the four significant maps of interest presented in Figs 4B and 4C.

### Supplementary Tables

|  | Baseline | 1 year | 3 years | 10 years |
| --- | --- | --- | --- | --- |
| Age inclusion | 30.08 ± 6.82 |  |  |  |
| Years of Education | 10.12 ± 2.55 |  |  |  |
| Global Cognitive Functioning Attention | 0.07 ± 1.02 | 0.2 ± 1.22 | 0.18 ± 0.7 | 0.29 ± 0.58 |
| Global Cognitive Functioning average | -0.34 ± 0.65 | 0.24 ± 0.67 | 0.17 ± 0.57 | 0.19 ± 0.49 |
| Global Cognitive Functioning Executive Function | -0.18 ± 1.14 | 0.24 ± 0.71 | 0.24 ± 0.66 | 0.04 ± 0.63 |
| Global Cognitive Functioning Motor Dexterity | -0.31 ± 1.14 | 0.17 ± 0.68 | 0.08 ± 1.11 | 0.51 ± 0.74 |
| Global Cognitive Functioning Processing Speed | -0.3 ± 0.98 | 0.29 ± 0.56 | 0.28 ± 0.79 | 0.38 ± 0.89 |
| Global Cognitive Functioning Verbal Memory | -1.67 ± 1.45 | -0.5 ± 1.18 | -0.6 ± 1.19 | -0.72 ± 1.17 |
| Global Cognitive Functioning Visual Memory | 0.09 ± 1.02 | 0.67 ± 0.89 | 0.89 ± 0.72 | 0.6 ± 0.65 |
| Global Cognitive Functioning Working Memory | -0.07 ± 0.88 | 0.62 ± 1.04 | 0.12 ± 0.86 | 0.2 ± 1.12 |

**Table S1.** Demographic and cognitive data for male control participants (mean ± standard deviation.

|  | Baseline | 1 year | 3 years | 10 years |
| --- | --- | --- | --- | --- |
| Age inclusion | 29.83 ± 8.16 |  |  |  |
| Years of Education | 11.64 ± 2.64 |  |  |  |
| Global Cognitive Functioning Attention | -0.11 ± 1.12 | -0.01 ± 1.22 | 0.29 ± 0.46 | -0.1 ± 1.16 |
| Global Cognitive Functioning average | -0.11 ± 0.55 | -0.01 ± 0.6 | 0.15 ± 0.44 | 0.12 ± 0.46 |
| Global Cognitive Functioning Executive Function | 0.18 ± 0.76 | 0.36 ± 0.59 | 0.21 ± 0.49 | 0.12 ± 0.59 |
| Global Cognitive Functioning Motor Dexterity | 0.28 ± 0.8 | 0.47 ± 0.61 | 0.8 ± 0.53 | 0.5 ± 0.96 |
| Global Cognitive Functioning Processing Speed | 0.33 ± 1.03 | 0.59 ± 0.85 | 0.52 ± 1.13 | 0.87 ± 0.77 |
| Global Cognitive Functioning Verbal Memory | -0.89 ± 1.05 | -1.08 ± 1.46 | -0.79 ± 0.89 | -0.61 ± 0.93 |
| Global Cognitive Functioning Visual Memory | -0.32 ± 1.01 | -0.19 ± 1.04 | 0.16 ± 0.53 | 0.04 ± 0.69 |
| Global Cognitive Functioning Working Memory | -0.21 ± 0.94 | -0.22 ± 0.65 | -0.15 ± 0.86 | -0.02 ± 0.79 |

**Table S2.** Demographic and cognitive data for female control participants (mean ± standard deviation.

|  | Baseline | 1 year | 3 years | 10 years |
| --- | --- | --- | --- | --- |
| Age inclusion | 27.62 ± 7.49 |  |  |  |
| Age of onset | 26.86 ± 7.5 |  |  |  |
| Years of Education | 9.59 ± 3.03 |  |  |  |
| BPRS depression | 2.4 ± 0.89 | 1.27 ± 0.6 | 1.26 ± 0.49 | 1.21 ± 0.38 |
| BPRS Disorientation | 3.16 ± 1.74 | 1.95 ± 1.3 | 1.72 ± 1.23 | 1.75 ± 0.96 |
| BPRS Mania | 2.56 ± 0.94 | 1.31 ± 0.51 | 1.26 ± 0.53 | 1.15 ± 0.27 |
| BPRS Negative | 1.79 ± 0.85 | 1.37 ± 0.52 | 1.38 ± 0.73 | 1.24 ± 0.41 |
| BPRS Positive | 3.33 ± 0.72 | 1.42 ± 0.7 | 1.23 ± 0.51 | 1.2 ± 0.4 |
| BPRS Total | 64.15 ± 14.06 | 33.55 ± 12.12 | 31.45 ± 11.56 | 29.81 ± 7.33 |
| Global Cognitive Functioning Attention | -1.86 ± 3.67 | -1.75 ± 4.63 | -1.3 ± 3.29 | -1 ± 2.41 |
| Global Cognitive Functioning average | -1.33 ± 1.15 | -0.98 ± 1.41 | -0.95 ± 0.95 | -1.03 ± 1.15 |
| Global Cognitive Functioning Executive Function | -1.07 ± 1.86 | -0.64 ± 1.45 | -0.76 ± 1.52 | -1.02 ± 2.32 |
| Global Cognitive Functioning Motor Dexterity | -1.27 ± 2.6 | -0.94 ± 3.74 | -0.67 ± 1.41 | -1.12 ± 2.25 |
| Global Cognitive Functioning Processing Speed | -1.69 ± 1.06 | -1.17 ± 1.3 | -1.25 ± 1.08 | -0.64 ± 1 |
| Global Cognitive Functioning Verbal Memory | -2.5 ± 1.32 | -2.19 ± 1.5 | -2.22 ± 1.4 | -2.33 ± 1.29 |
| Global Cognitive Functioning Visual Memory | -0.44 ± 1 | 0.09 ± 1.05 | -0.13 ± 0.99 | -0.6 ± 0.84 |
| Global Cognitive Functioning Working Memory | -0.43 ± 0.78 | -0.5 ± 0.8 | -0.32 ± 0.78 | -0.47 ± 0.87 |
| SANS Global Rating of Affective Flattening | 1.24 ± 1.49 | 1.3 ± 1.48 | 0.95 ± 1.39 | 1.13 ± 1.24 |
| SANS Global Rating of Alogia | 0.84 ± 1.46 | 0.67 ± 1.13 | 0.78 ± 1.28 | 0.72 ± 1.12 |
| SANS Global Rating of Anhedonia Asociality | 1.53 ± 1.8 | 1.48 ± 1.68 | 1.14 ± 1.71 | 1.24 ± 1.67 |
| SANS Global Rating of Attention | 1.85 ± 1.96 | 0.45 ± 1.12 | 0.57 ± 1.28 | 0.17 ± 0.5 |
| SANS Global Rating of Avolition Apathy | 1.07 ± 1.69 | 1.39 ± 1.7 | 1.09 ± 1.61 | 0.81 ± 1.37 |
| SANS Total | 1.17 ± 1.31 | 1.21 ± 1.3 | 0.99 ± 1.34 | 0.98 ± 1.18 |
| SAPS Global rating of bizarre behaviour | 4.18 ± 1.42 | 0.45 ± 1.09 | 0.33 ± 0.99 | 0.15 ± 0.6 |
| SAPS Global rating of delusions | 4.87 ± 0.52 | 1.09 ± 1.62 | 0.66 ± 1.3 | 0.69 ± 1.15 |
| SAPS Global rating of hallucinations | 2.65 ± 2.32 | 0.31 ± 1.02 | 0.37 ± 1.09 | 0.22 ± 0.77 |
| SAPS Global rating of positive formal thought disorder | 0.99 ± 1.7 | 0.09 ± 0.54 | 0.13 ± 0.59 | 0.06 ± 0.3 |
| SAPS Total | 3.17 ± 0.82 | 0.49 ± 0.77 | 0.37 ± 0.79 | 0.28 ± 0.54 |

**Table S3.** Demographic, cognitive and clinical data for male FEP participants (mean ± standard deviation.

|  | Baseline | 1 year | 3 years | 10 years |
| --- | --- | --- | --- | --- |
| Age inclusion | 32.94 ± 9.61 |  |  |  |
| Age of onset | 32.13 ± 9.52 |  |  |  |
| Years of Education | 11.26 ± 3.42 |  |  |  |
| BPRS Depression | 2.5 ± 0.95 | 1.31 ± 0.4 | 1.21 ± 0.3 | 1.48 ± 0.61 |
| BPRS Disorientation | 2.7 ± 1.73 | 1.34 ± 0.65 | 1.29 ± 0.59 | 1.5 ± 0.61 |
| BPRS Mania | 2.6 ± 1.1 | 1.13 ± 0.26 | 1.12 ± 0.19 | 1.17 ± 0.28 |
| BPRS Negative | 1.83 ± 0.87 | 1.19 ± 0.37 | 1.18 ± 0.35 | 1.21 ± 0.48 |
| BPRS Positive | 3.32 ± 0.79 | 1.06 ± 0.24 | 1.1 ± 0.27 | 1.15 ± 0.33 |
| BPRS Total | 64.07 ± 15.65 | 28.13 ± 5.31 | 27.77 ± 5.3 | 30.27 ± 6.99 |
| Global Cognitive Functioning Attention | -2.39 ± 3.88 | -2.36 ± 4.24 | -1.72 ± 3.59 | -1.87 ± 4.08 |
| Global Cognitive Functioning average | -1.27 ± 1.15 | -0.88 ± 1.33 | -0.77 ± 1.03 | -0.9 ± 1.1 |
| Global Cognitive Functioning Executive Function | -1.22 ± 2.08 | -0.91 ± 2.72 | -0.68 ± 1.68 | -0.6 ± 1.37 |
| Global Cognitive Functioning Motor Dexterity | -0.88 ± 1.91 | -0.08 ± 1.19 | -0.32 ± 1.23 | -0.62 ± 1.4 |
| Global Cognitive Functioning Processing Speed | -0.99 ± 1.07 | -0.43 ± 1.13 | -0.44 ± 1.27 | -0.41 ± 0.95 |
| Global Cognitive Functioning Verbal Memory | -2.11 ± 1.44 | -1.94 ± 1.36 | -1.46 ± 1.4 | -1.61 ± 1.37 |
| Global Cognitive Functioning Visual Memory | -0.69 ± 0.94 | -0.21 ± 1.25 | -0.49 ± 0.81 | -0.61 ± 0.86 |
| Global Cognitive Functioning Working Memory | -0.55 ± 1.02 | -0.21 ± 0.94 | -0.27 ± 0.78 | -0.56 ± 0.85 |
| SANS Global Rating of Affective Flattening | 0.79 ± 1.32 | 0.71 ± 1.14 | 0.45 ± 0.89 | 0.81 ± 1.29 |
| SANS Global Rating of Alogia | 0.63 ± 1.31 | 0.39 ± 1.03 | 0.26 ± 0.68 | 0.54 ± 1.07 |
| SANS Global Rating of Anhedonia Asociality | 0.98 ± 1.6 | 1.05 ± 1.54 | 0.66 ± 1.09 | 1.43 ± 1.66 |
| SANS Global Rating of Attention | 2.06 ± 1.92 | 0.45 ± 1.16 | 0.13 ± 0.34 | 0.41 ± 0.83 |
| SANS Global Rating of Avolition Apathy | 0.68 ± 1.38 | 0.74 ± 1.11 | 0.49 ± 0.93 | 0.76 ± 1.12 |
| SANS Total | 0.77 ± 1.14 | 0.72 ± 1.02 | 0.47 ± 0.74 | 0.89 ± 1.06 |
| SAPS Global rating of bizarre behaviour | 4.2 ± 1.29 | 0.05 ± 0.32 | 0.19 ± 0.62 | 0.14 ± 0.54 |
| SAPS Global rating of delusions | 4.75 ± 0.75 | 0.18 ± 0.8 | 0.4 ± 1.12 | 0.51 ± 1.22 |
| SAPS Global rating of hallucinations | 2.27 ± 2.29 | 0.11 ± 0.51 | 0.06 ± 0.3 | 0.19 ± 0.74 |
| SAPS Global rating of positive formal thought disorder | 1.25 ± 1.8 | 0 ± 0 | 0.02 ± 0.14 | 0.16 ± 0.69 |
| SAPS Total | 3.12 ± 0.92 | 0.09 ± 0.31 | 0.17 ± 0.41 | 0.25 ± 0.62 |

**Table S4.** Demographic, cognitive and clinical data for female FEP participants (mean ± standard deviation.

| DK area | Mesulam Area |
| --- | --- |
| inferiorparietal | heteromodal |
| middletemporal | heteromodal |
| parsorbitalis | heteromodal |
| parstriangularis | heteromodal |
| precuneus | heteromodal |
| rostralmiddlefrontal | heteromodal |
| superiorfrontal | heteromodal |
| superiorparietal | heteromodal |
| supramarginal | heteromodal |
| frontalpole | heteromodal |
| cuneus | idiotypic |
| lingual | idiotypic |
| paracentral | idiotypic |
| pericalcarine | idiotypic |
| postcentral | idiotypic |
| precentral | idiotypic |
| caudalanteriorcingulate | paralimbic |
| entorhinal | paralimbic |
| isthmuscingulate | paralimbic |
| lateralorbitofrontal | paralimbic |
| medialorbitofrontal | paralimbic |
| parahippocampal | paralimbic |
| posteriorcingulate | paralimbic |
| rostralanteriorcingulate | paralimbic |
| temporalpole | paralimbic |
| insula | paralimbic |
| bankssts | unimodal |
| caudalmiddlefrontal | unimodal |
| fusiform | unimodal |
| inferiortemporal | unimodal |
| lateraloccipital | unimodal |
| parsopercularis | unimodal |
| superiortemporal | unimodal |
| transversetemporal | unimodal |

**Table S5.** Spatial overlap between Desikan-Killiany and Mesulam zones.

| **Abbreviation** | **Neurobiological feature** | **Type** |
| --- | --- | --- |
| 5-HT_1A_ | Serotonin receptor | Neurotransmitter |
| 5-HT1_B_ | Serotonin receptor | Neurotransmitter |
| 5-HT_2A_ | Serotonin receptor | Neurotransmitter |
| 5-HT_4_ | Serotonin receptor | Neurotransmitter |
| 5-HT_6_ | Serotonin receptor | Neurotransmitter |
| 5-HTT | Serotonin transporter | Neurotransmitter |
| H_3_ | Histamine receptor | Neurotransmitter |
| D_1_ | Dopamine receptor | Neurotransmitter |
| D_2_ | Dopamine receptor | Neurotransmitter |
| DAT | Dopamine transporter | Neurotransmitter |
| NET | Norepinephrine transporter | Neurotransmitter |
| α_4_β_2_ | Acetylcholine receptor | Neurotransmitter |
| M_1_ | Acetylcholine receptor | Neurotransmitter |
| VAChT | Acetylcholine transporter | Neurotransmitter |
| CB_1_ | Cannabinoid receptor | Neurotransmitter |
| MOR | Opioid receptor | Neurotransmitter |
| mGluR_5_ | Glutamate receptor | Neurotransmitter |
| NMDA | Glutamate receptor | Neurotransmitter |
| GABA | GABA receptor | Neurotransmitter |
| Astro | Astrocytes | Cell type |
| Endo | Endothelial cells | Cell type |
| Micro | Microglia | Cell type |
| Oligo | Oligodendrocytes | Cell type |
| OPC | Oligodendrocytes precursors | Cell type |
| Neuro-Ex | Excitatory neurons | Cell type |
| Neuro-In | Inhibitory neurons | Cell type |
| Layer I | Layer I | Layer thickness |
| Layer II | Layer II | Layer thickness |
| Layer III | Layer III | Layer thickness |
| Layer IV | Layer IV | Layer thickness |
| Layer V | Layer V | Layer thickness |
| Layer VI | Layer VI | Layer thickness |
| Myelin | Myelin | Microstructure |
| Thickness | Cortical thickness | Microstructure |
| Gene PC1 | Gene expression PC1 | Microstructure |
| Neurotransmitter PC1 | Neurotransmitter PC1 | Microstructure |
| Synapse density | Synapse density | Microstructure |
| Evolutionary exp. | Evolutionary expansion | Cortical expansion |
| Developmental exp. | Developmental expansion | Cortical expansion |
| Scaling PNC | Allometric scaling from Philadelphia Neurodevelopmental Cohort | Cortical expansion |
| Scaling NIH | Allometric scaling from National Institutes of Health | Cortical expansion |
| CBF | Cerebral blood flow | Metabolism |
| CBV | Cerebral blood volume | Metabolism |
| CMRO_2_ | Oxygen metabolism | Metabolism |
| CMRGlu | Glucose metabolism | Metabolism |
| Glycolytic index | Glycolytic index | Metabolism |

**Table S6.** Molecular, microarchitectural and metabolic features included in the neurobiological mapping.

|  | | **Total grey matter volume centile** | **Total white matter volume centile** |
| --- | --- | --- | --- |
| **Intercept** | **t**-**value** | 29.615 | 28.34 |
|  | **p-value** | < 10^-15^ | < 10^-15^ |
| **Diagnosis** | **t-value** | -6.20 | -1.67 |
|  | **p-value** | < 10^-8^ | 0.095 |
| **Age of Inclusion** | **t-value** | 4.52 | 3.75 |
|  | **p-value** | <10^-5^ | <10^-4^ |
| **Sex** | **t-value** | -2.27 | 3.75 |
|  | **p-value** | 0.023 | <10^-4^ |
| **Etiv** | **t-value** | 18.03 | 16.47 |
|  | **p-value** | < 10^-15^ | < 10^-15^ |

**Table S7.** Linear multiple regression of total grey and white matter volume centiles (baseline data only). Statistically significant values are represented in red (p-value<0.05).

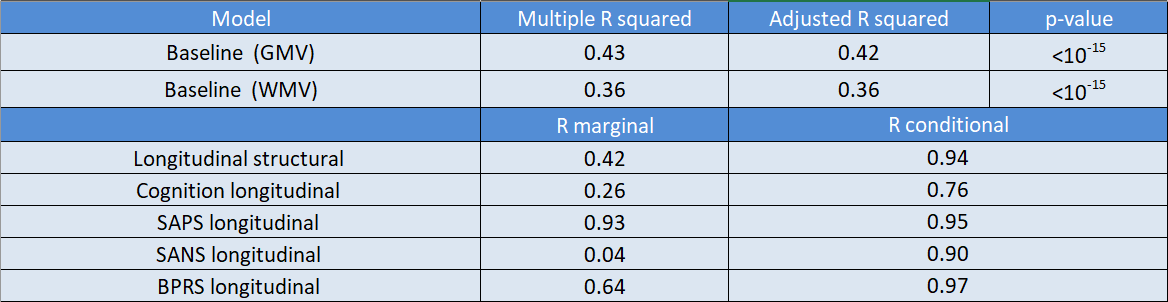

**Table S8.** Fitting estimation for global models.

|  | | **Total grey matter volume centile** | **Total white matter volume centile** |
| --- | --- | --- | --- |
| **Intercept** | **t**-**value** | 29.48 | 28.94 |
|  | **p-value** | < 10^-15^ | <10^-15^ |
| **Diagnosis** | **t-value** | -4.55 | -1.71 |
|  | **p-value** | <10^-5^ | 0.087 |
| **Age of Inclusion** | **t-value** | 4.49 | 3.71 |
|  | **p-value** | <10^-5^ | <10^-3^ |
| **Sex** | **t-value** | -2.64 | -1.94 |
|  | **p-value** | 0.008 | 0.05 |
| **Etiv** | **t-value** | 19.28 | 17.35 |
|  | **p-value** | <10^-15^ | <10^-15^ |
| **Medication** | **t-value** | -3.31 | 0.57 |
|  | **p-value** | <10^-3^ | 0.57 |
| **Time** | **t-value** | -0.8 | 1.60 |
|  | **p-Value** | 0.42 | 0.087 |
| **Time: FEP** | **t-value** | 4.02 | -2.45 |
|  | **p-value** | <10^-4^ | 0.015 |
| **Medication : Time** | **t-value** | -5.04 | -1.25 |
|  | **p-value** | <10^-6^ | 0.21 |

**Table S9**. Linear mixed regression of total grey and white matter volume centiles (longitudinal data). Significant values in red (p-value <0.05).

| **Principal Components Analysis of cognitive subscales** | **PC1** | **PC2** | **PC3** |
| --- | --- | --- | --- |
| **Global Cognitive Functioning Verbal Memory** | 0.37 | -0.4 | -0.58 |
| **Global Cognitive Functioning Visual Memory** | 0.38 | -0.23 | 0.04 |
| **Global Cognitive Functioning Processing Speed** | 0.41 | 0.03 | -0.42 |
| **Global Cognitive Functioning Working Memory** | 0.34 | -0.53 | 0.53 |
| **Global Cognitive Functioning Executive Function** | 0.40 | 0.09 | 0.36 |
| **Global Cognitive Functioning Motor Dexterity** | 0.37 | 0.54 | -0.12 |
| **Global Cognitive Functioning Attention** | 0.37 | 0.45 | 0.24 |

**Table S10.** Principal component analysis of cognitive subscales data.

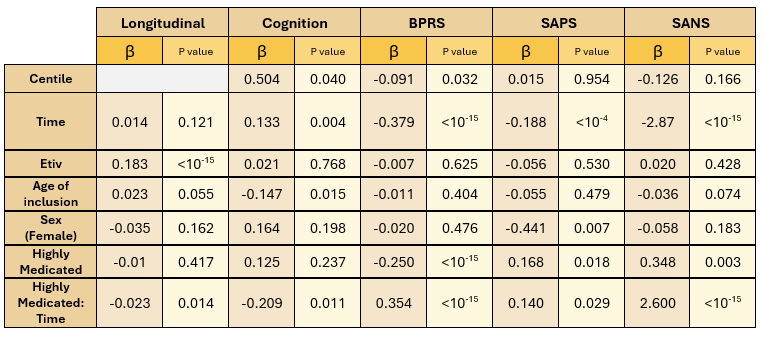

**Table S11.** LMMs after categorizing patients into low (CPZ≤300) and high medication groups (CPZ>300).

| **Clinical scale** | **Assessment** | **FEP cannabis users** | **FEP cannabis non-users** | **p Value** |
| --- | --- | --- | --- | --- |
| **Average** | 1 | -1.24 ± 0.86 | -1.35 ± 1.30 | 0.5391 |
| **Cognitive** | 2 | -0.77 ± 0.95 | -0.98 ± 1.44 | 0.5956 |
| **Functioning** | 3 | -0.89 ± 1.16 | -0.88 ± 0.95 | 0.9754 |
|  | 10 | -0.91 ± 1.35 | -0.98 ± 1.12 | 0.8873 |
|  | 1 | 3.16 ± 0.89 | 3.14 ± 0.84 | 0.8131 |
| **SAPS** | 2 | 0.86 ± 0.87 | 0.23 ± 0.56 | 0.4911 |
|  | 3 | 0.67 ± 1.24 | 0.24 ± 0.55 | 0.4808 |
|  | 10 | 0.57 ± 0.62 | 0.24 ± 0.56 | 0.4907 |
|  | 1 | 0.99 ± 1.20 | 1.03 ± 1.30 | 0.7624 |
| **SANS** | 2 | 1.01 ± 1.16 | 1.04 ± 1.25 | 0.9346 |
|  | 3 | 0.97 ± 1.25 | 0.78 ± 1.18 | 0.5693 |
|  | 10 | 0.96 ± 0.88 | 0.94 ± 1.15 | 0.9336 |
|  | 1 | 63.55 ± 13.69 | 64.48 ± 15.31 | 0.6189 |
| **BPRS** | 2 | 35.74 ± 12.92 | 30.67 ± 9.73 | 0.4879 |
|  | 3 | 36.61 ± 17.99 | 29.21 ± 7.87 | 0.473 |
|  | 10 | 32.57 ± 8.12 | 29.79 ± 7.08 | 0.4995 |

**Table S12.** Mean (± standard deviation) clinical scores stratified for cannabis users and non-users. P-values were computed using the Wilcoxon signed-rank test.

| Variable | Assessment transition | N dropout | N continue | Median  dropout | Median  continue | P Value | P Value  Adj FDR |
| --- | --- | --- | --- | --- | --- | --- | --- |
| Cognition | 1🡪2 | 211 | 91 | -1.00 | -1.09 | 0.448 | 0.672 |
|  | 2🡪3 | 21 | 61 | -0.25 | -0.75 | 0.048 | 0.144 |
|  | 3🡪10 | 73 | 48 | -0.72 | -0.74 | 0.745 | 0.745 |
| SAPS | 1🡪2 | 248 | 103 | 3.50 | 3.00 | 0.540 | 0.614 |
|  | 2🡪3 | 26 | 79 | 0.00 | 0.00 | 0.330 | 0.614 |
|  | 3🡪10 | 91 | 55 | 0.00 | 0.00 | 0.614 | 0.614 |
| SANS | 1🡪2 | 248 | 103 | 0.25 | 0.75 | 0.069 | 0.207 |
|  | 2🡪3 | 26 | 79 | 0.50 | 0.50 | 0.562 | 0.772 |
|  | 3🡪10 | 91 | 55 | 0.25 | 0.25 | 0.772 | 0.772 |
| BPRS | 1🡪2 | 248 | 103 | 64.00 | 60.00 | 0.072 | 0.216 |
|  | 2🡪3 | 26 | 79 | 27.00 | 27.00 | 0.703 | 0.703 |
|  | 3🡪10 | 90 | 55 | 26.00 | 27.00 | 0.590 | 0.703 |

**Table S13.** Comparison of clinical characteristics between participants who continued and those who dropped out at subsequent assessments. Results are presented for transitions from assessment 1 to 2, 2 to 3, and 3 to 10. The table includes median values, sample sizes, and Wilcoxon rank-sum p-values for cognition, SAPS, SANS, and BPRS. P-values were corrected for multiple testing using the False Discovery Rate (FDR) adjustment.
